# Negative BOLD Responses Surpass Positive Responses in Task Specificity, Reflecting Neural Reconfigurations Better Than Functional Connectivity

**DOI:** 10.1101/2024.11.20.24317658

**Authors:** Saman Gholipour Picha, Seyed Hani Hojjati, Siddharth Nayak, Sindy Ozoria, Peter Chernek, Jenseric Calimag, Bardiya Ghaderi Yazdi, Qolamreza R. Razlighi

## Abstract

**Objective:** To investigate whether the Negative BOLD Response (NBR) is more task-specific than the Positive BOLD Response (PBR) during cognitive tasks and to determine whether task-evoked activity reflects brain reconfigurations during different tasks better than functional connectivity.

**Methods:** Functional Magnetic Resonance Imaging (fMRI) data were collected from 214 participants under 50 years old (152 in Dataset 1 and 62 in Dataset 2) performing twelve cognitive tasks spanning vocabulary, speed of processing, fluid reasoning, and memory domains. Data analysis included subject-level and group-level analyses, focusing on comparing the spatial patterns and task specificity of NBR and PBR through similarity measures using Dice coefficients. Additionally, functional connectivity was assessed using the Multi-session Hierarchical Bayesian Model (MS-HBM) to evaluate its sensitivity to task-induced brain reconfigurations compared to task-evoked activity.

**Results:** NBR demonstrated significantly greater task specificity compared to PBR across all cognitive tasks, with lower mean Dice coefficients for NBR maps (mean: 0.44, SD: 0.13) than for PBR maps (mean: 0.67, SD: 0.09; t(65) = 18.38, p < 0.001). Functional connectivity analyses indicated that the default mode network (DMN) remained stable across tasks, suggesting that task-evoked activity reflects task-specific brain reconfigurations better than functional connectivity.

**Conclusion:** The findings confirm that NBR is inherently more task-specific than PBR and that task-evoked activity provides a more sensitive measure of task-specific neural reconfigurations than functional connectivity. This enhances our understanding of the neural mechanisms underlying cognitive processes and highlights the importance of considering NBR in cognitive neuroscience research.

## 1 Introduction

The ability of the human brain to perform a vast array of cognitive tasks relies on complex neural mechanisms that are still not fully understood. Functional Magnetic Resonance Imaging (fMRI) has been pivotal in unraveling these mechanisms by allowing researchers to non-invasively map brain activity associated with various cognitive processes. Central to fMRI studies is the Blood Oxygen Level Dependent (BOLD) signal, reflecting changes in blood oxygenation linked to neural activity (Ogawa et al., 1990).

The BOLD signal comprises both Positive BOLD Responses (PBR), which are increases in signal intensity associated with enhanced neural activity, and Negative BOLD Responses (NBR), decreases in signal intensity often linked to neural deactivation (Shmuel et al., 2006). While PBRs have been extensively studied over the past decades, NBRs have received comparatively less attention despite accumulating evidence suggesting their significant role in cognitive functions (Logothetis et al., 2001).

NBRs are often observed in regions that are part of the Default Mode Network (DMN), a set of brain areas showing decreased activity during externally focused tasks and increased activity during rest or internally directed thought (Raichle et al., 2001)(Buckner et al., 2008). The DMN includes regions such as the medial prefrontal cortex, posterior cingulate cortex, and inferior parietal lobule. Deactivation of the DMN (manifested as NBR) during task performance is thought to reflect the suspension of self-referential and introspective processes to facilitate attention to external stimuli (Anticevic et al., 2012)(Whitfield-Gabrieli & Ford, 2012).

Understanding the task specificity of NBR is crucial because it may provide insights into how the brain reallocates resources to support different cognitive demands. Previous studies have suggested that NBR may be more sensitive to task demands than PBR, indicating potential for greater task specificity (Newton et al., 2011). Despite accumulating evidence of NBR’s significance, systematic investigations directly comparing the task specificity of NBR and PBR across a wide range of cognitive tasks are scarce. This gap hampers a comprehensive understanding of how these responses contribute differently to cognitive processing.

Moreover, functional connectivity analyses have revealed that the brain’s functional networks can reorganize in response to task demands (Cole et al., 2014). Functional connectivity refers to the temporal correlation between spatially remote neurophysiological events, reflecting how different brain regions communicate during rest or task performance (Friston, 2011). It remains to be determined whether task-evoked activity (NBR and PBR) or functional connectivity better reflects the brain’s reconfigurations during different cognitive tasks.

Addressing these objectives has significant implications for both basic neuroscience and clinical applications. If NBR is inherently more task-specific than PBR and task-evoked activity reflects task-induced brain reconfigurations better than functional connectivity, NBR could serve as a more sensitive biomarker for assessing cognitive function and dysfunction. This has potential applications in diagnosing and monitoring neurological and psychiatric conditions where DMN activity is altered, such as Alzheimer’s disease, depression, and schizophrenia (Broyd et al., 2009)(Anticevic et al., 2012)(Whitfield-Gabrieli & Ford, 2012).

Furthermore, understanding the relative effectiveness of task-evoked activity and functional connectivity in reflecting task-specific brain reconfigurations can enhance models of brain network dynamics, contributing to theories of cognitive control, attentional processes, and the neural basis of cognition (Menon, 2011).

### 1.1 Objectives of the Study

Given these considerations, the primary objectives of this study are to determine whether the Negative BOLD Response exhibits greater task specificity than the Positive BOLD Response during cognitive tasks, and to assess whether task-evoked activity reflects brain reconfigurations during different tasks better than functional connectivity. By analyzing a broad spectrum of cognitive tasks across different domains, we aim to systematically compare the spatial patterns of NBR and PBR to assess their specificity to task demands. Additionally, we seek to determine which measure—task-evoked activity (NBR and PBR) or functional connectivity—more accurately captures task-specific neural dynamics.

## 2 Methods

### 2.1 Participants

Participants were recruited through random-market mailing within a 10-mile radius of Columbia University Irving Medical Center (Dataset 1) and Weill Cornell Medical Center (Dataset 2). The inclusion criteria were designed to ensure a homogeneous sample of healthy adults, minimizing potential confounding variables. Participants in Dataset 1 were aged between 19 and 49 years, while those in Dataset 2 were aged between 19 and 39 years. All participants were right-handed, as assessed by the Edinburgh Handedness Inventory, to control for hemispheric dominance effects. They had normal or corrected-to-normal vision and were fluent English speakers to ensure comprehension of task instructions.

Exclusion criteria included a history of neurological or psychiatric disorders, such as head injury, epilepsy, or major depression, to avoid confounding factors related to brain function. Participants with contraindications to MRI scanning, such as metal implants or claustrophobia, were also excluded.

In total, 152 participants (90 females, 62 males) were included in Dataset 1, and 62 participants (30 females, 32 males) were included in Dataset 2. Detailed demographic information, including age distribution and gender breakdown for each task, is provided in Table 1.

**Table 1.**
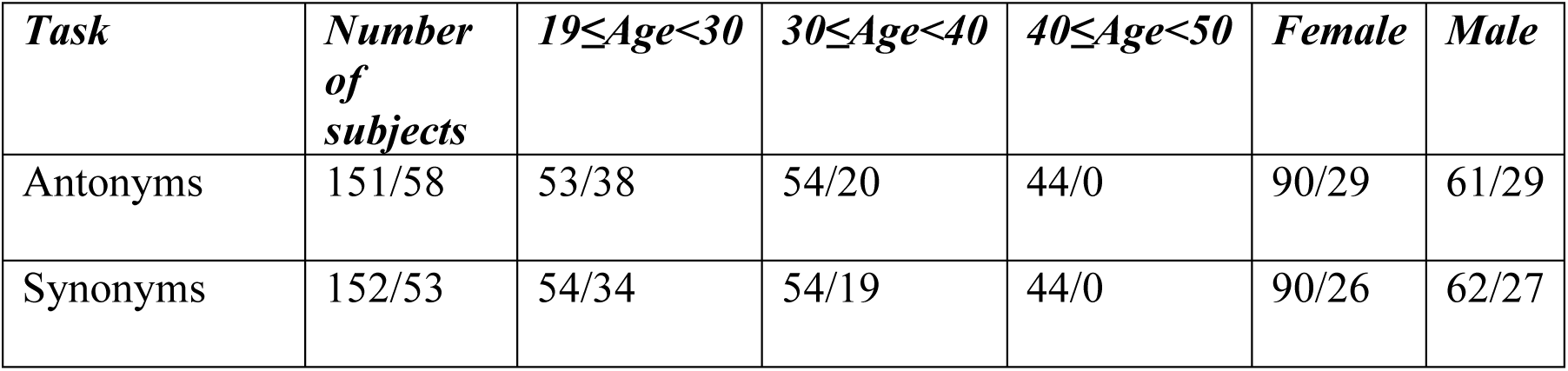
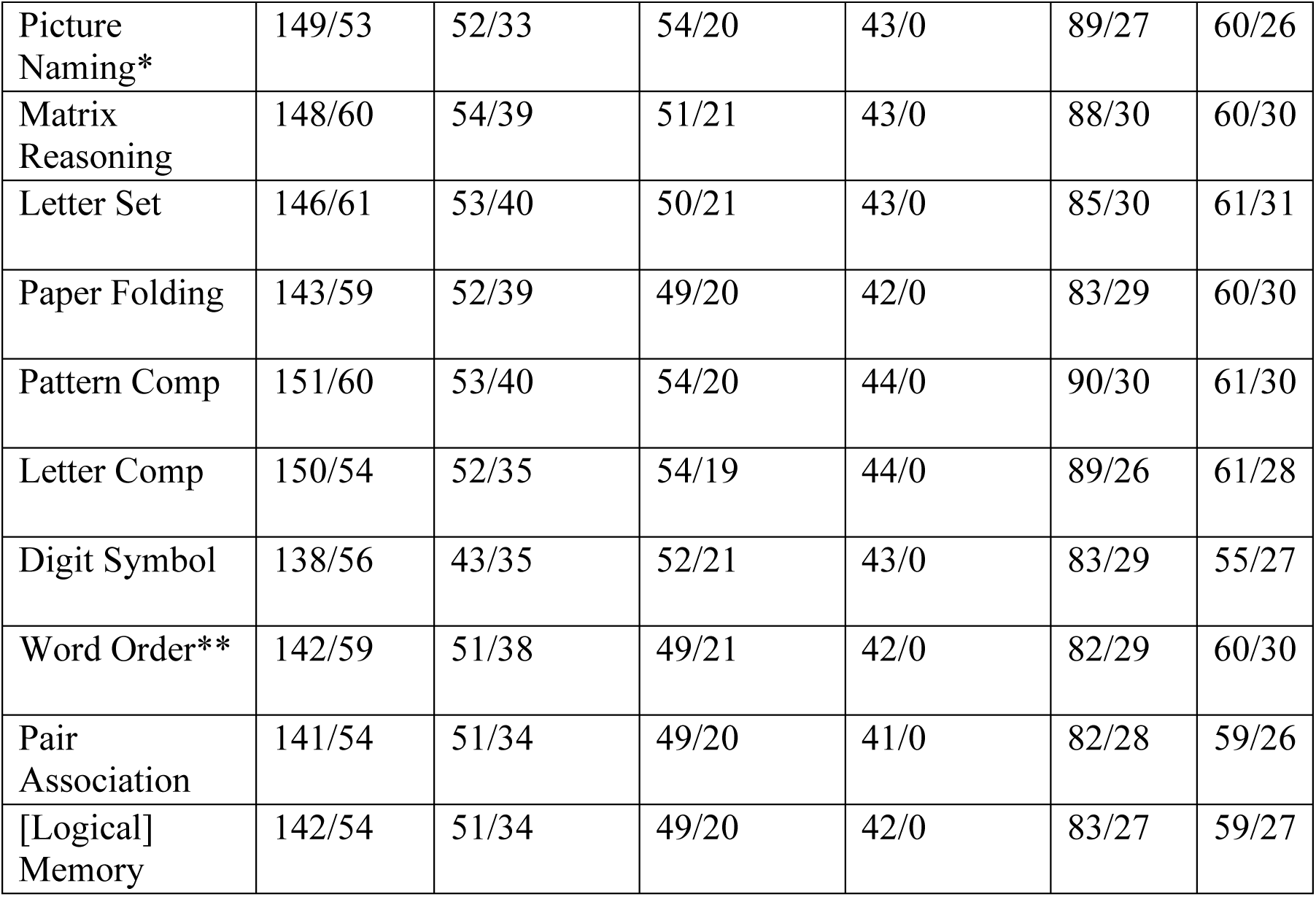
Demographics for two datasets; first numbers are the number of subjects for dataset 1. There are some minor differences between the tasks among different datasets. * Picture Vocabulary in dataset 2 ** Word Memorization in dataset 2.

| **Task** | **Number of subjects** | **19≤Age<30** | **30≤Age<40** | **40≤Age<50** | **Female** | **Male** |
| --- | --- | --- | --- | --- | --- | --- |
| Antonyms | 151/58 | 53/38 | 54/20 | 44/0 | 90/29 | 61/29 |
| Synonyms | 152/53 | 54/34 | 54/19 | 44/0 | 90/26 | 62/27 |
| Picture Naming* | 149/53 | 52/33 | 54/20 | 43/0 | 89/27 | 60/26 |
| Matrix Reasoning | 148/60 | 54/39 | 51/21 | 43/0 | 88/30 | 60/30 |
| Letter Set | 146/61 | 53/40 | 50/21 | 43/0 | 85/30 | 61/31 |
| Paper Folding | 143/59 | 52/39 | 49/20 | 42/0 | 83/29 | 60/30 |
| Pattern Comp | 151/60 | 53/40 | 54/20 | 44/0 | 90/30 | 61/30 |
| Letter Comp | 150/54 | 52/35 | 54/19 | 44/0 | 89/26 | 61/28 |
| Digit Symbol | 138/56 | 43/35 | 52/21 | 43/0 | 83/29 | 55/27 |
| Word Order** | 142/59 | 51/38 | 49/21 | 42/0 | 82/29 | 60/30 |
| Pair Association | 141/54 | 51/34 | 49/20 | 41/0 | 82/28 | 59/26 |
| [Logical] Memory | 142/54 | 51/34 | 49/20 | 42/0 | 83/27 | 59/27 |

All participants provided written informed consent approved by the Institutional Review Boards of both institutions. They were compensated for their participation and informed of their right to withdraw from the study at any point without penalty.

### 2.2 Data Acquisition Protocols

High-resolution structural and functional MRI data were acquired using 3.0 Tesla scanners at both institutions, ensuring high spatial resolution and optimal signal-to-noise ratios.

For Dataset 1, imaging was performed on a Philips Achieva 3.0 Tesla scanner using an 8-channel receive-only head coil. Functional images were acquired using a T2*-weighted echo-planar imaging (EPI) sequence sensitive to BOLD contrast. The imaging parameters were as follows: repetition time (TR) = 2000 ms, echo time (TE) = 20 ms, flip angle = 72°, field of view (FOV) = 224 × 224 mm, matrix size = 112 × 112, in-plane resolution = 2 × 2 mm, slice thickness = 4 mm with no gap, and 33 axial slices covering the entire brain. The duration of functional scans varied by task, ranging from 198 seconds for shorter tasks to 860 seconds for longer tasks.

Structural images for Dataset 1 were acquired using a T1-weighted magnetization-prepared rapid gradient-echo (MPRAGE) sequence with TR = 6.6 ms, TE = 2.98 ms, flip angle = 8°, FOV = 256 × 256 mm, matrix size = 256 × 256, voxel size = 1 × 1 × 1 mm³, and 180 axial slices.

For Dataset 2, imaging was performed on a Siemens Magnetom Prisma 3.0 Tesla scanner using a 64-channel receive-only head coil. Functional images were acquired using a T2*-weighted multiband EPI sequence to reduce TR and increase temporal resolution. The parameters were: TR = 1008 ms, TE = 37 ms, flip angle = 52°, FOV = 208 × 208 mm, matrix size = 104 × 104, in-plane resolution = 2 × 2 mm, slice thickness = 2 mm with no gap, 72 axial slices with a multiband factor of 6, capturing the entire brain. Each functional scan lasted 10 minutes and 5 seconds per task, totaling 600 volumes.

Structural images for Dataset 2 were acquired using a T1-weighted MPRAGE sequence with TR = 2400 ms, TE = 2.96 ms, flip angle = 9°, FOV = 256 × 256 mm, matrix size = 512 × 512, voxel size = 0.5 × 0.5 × 0.5 mm³, and 416 sagittal slices.

These imaging parameters were optimized to balance spatial resolution, signal-to-noise ratio, and scan duration, facilitating precise localization of brain activity while maintaining participant comfort.

### 2.3 Cognitive Tasks

Twelve cognitive tasks were administered during fMRI scanning, each designed to engage specific cognitive domains. Tasks were adapted from standardized neuropsychological assessments to ensure validity and reliability.

#### 2.3.1 Vocabulary Tasks

Assessed language comprehension, verbal reasoning, and crystallized intelligence. **Antonyms**. Participants were presented with a target word in uppercase at the top of the screen and four numbered choice words below. They were instructed to select the word that was most nearly opposite in meaning to the target word. **Synonyms**. Similar format to the Antonyms task, but participants selected the word most similar in meaning to the target word. **Picture Naming** (Dataset 1 only). Participants viewed images of common objects or scenes and were instructed to name them aloud. **Picture Vocabulary** (Dataset 2 only). Participants were shown four images in the screen’s corners and a word in the center. They selected the image that best matched the word.

#### 2.3.2 Speed of Processing Tasks

Evaluated the ability to process simple or complex information rapidly and accurately. **Digit Symbol**. A code table pairing digits (1–9) with unique symbols was displayed. Participants saw a sequence of symbols and had to identify the corresponding digit from the code table. **Letter Comparison**. Participants were shown pairs of letter strings and determined whether they were identical or differed. **Pattern Comparison**. Similar to Letter Comparison but with geometric patterns instead of letters. Patterns varied in complexity (lines, shapes).

Figure 1 shows the tasks paradigm. The tasks specifics are provided in Table S1 and Table S2. The schematic of both vocabulary and speed of processing tasks from dataset 1 and dataset 2 can be found in Figure 1.a and Figure 1.d respectively. Task design and timing can be found in Supp1.

**Figure 1.**
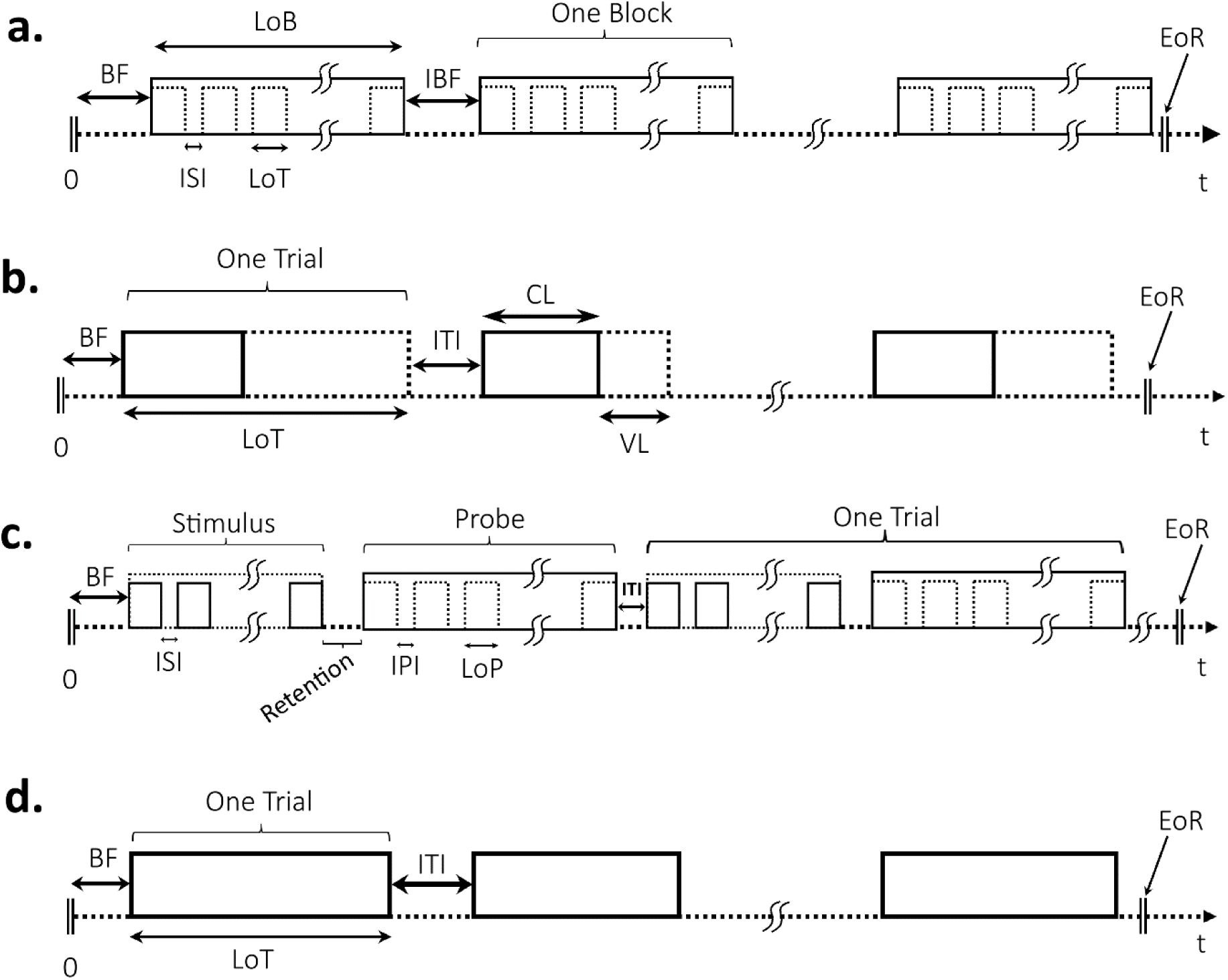
Tasks paradigm for (a) Vocabulary and Speed of Processing in dataset 1 (b) Fluid Reasoning for dataset 1 (CL is 11 s and VL is 74 s at max) (c) Memory for both datasets (Retention is 10 s) and (d) Vocabulary, Speed of Processing, and Fluid Reasoning for dataset 2 (EoR is 10’ 5”). BF: beginning fixation; EoR: end of run; NoS: number of stimuli; NoP: number of probe; TpB: trial per block; TpR: trial per run; LoT: length of trial; LoB: length of block; LoS: length of stimulus; LoP: length of probe; ISI: inter stimuli interval; ITI: inter trial interval; IPI: inter probes interval; RP: retention period; CL: compulsory length; VL: variable length.

#### 2.3.3 Fluid Reasoning Tasks

Measured problem-solving abilities and logical reasoning with novel information. **Paper Folding**. Illustrated sequences of paper folding and hole punching. Participants predicted the resulting pattern of holes when the paper was unfolded. **Matrix Reasoning**. Participants completed visual patterns in matrices by selecting the correct missing piece from multiple options. **Letter Sets**. Participants were presented with groups of letter sets and had to identify the set that did not belong based on underlying rules. The schematic of fluid reasoning tasks from dataset 1 and dataset 2 can be found in Figure 1.b and Figure 1.d respectively.

#### 2.3.4 Memory Tasks

Assessed various aspects of memory function, including encoding, storage, and retrieval. **Logical Memory**. Participants read short stories and were later asked to recall specific details. **Word Order** (Dataset 1 only). Participants viewed a list of words presented sequentially and later identified which word followed a given probe word. **Word Memorization** (Dataset 2 only). Participants viewed twelve words, each displayed individually with brief pauses between them. They were instructed to remember the words and later selected one of the words from multiple choices which was among the twelve words. **Paired Associates**. Participants learned pairs of unrelated words and were tested on their ability to recall the second word when presented with the first. The schematic of memory tasks from both datasets can be found in Figure 1.c.

Tasks were designed with fixed timings and consistent formats to control for extraneous variables affecting cognitive processing. Stimulus presentation durations, inter-stimulus intervals, and response recording methods were standardized across participants. Task order and stimulus presentation were randomized to prevent order effects. Detailed descriptions of task paradigms, and timing parameters are provided in the Supplementary Materials.

### 2.4 Data Preprocessing

Structural data were processed using FreeSurfer (version 7.1) for cortical reconstruction and volumetric segmentation, providing detailed anatomical information for subsequent analyses.

Functional data preprocessing differed between the two datasets due to differences in acquisition protocols.

For Dataset 1, functional MRI data were preprocessed using FSL (FMRIB Software Library) following standard procedures, including motion correction, slice timing correction, spatial smoothing, temporal filtering, and normalization to MNI space.

For Dataset 2, an in-house preprocessing pipeline was implemented to optimize data quality and address specific challenges associated with multiband echo-planar imaging (EPI) sequences. The pipeline, depicted in Figure S1, began with TopUp distortion correction using FSL’s TopUp tool to correct susceptibility-induced distortions that are common in EPI data. Following this, motion correction and realignment were performed using FSL’s MCFLIRT algorithm with the single-band reference (SBRef) image to ensure accurate alignment of functional images over time.

Slice timing correction was applied to adjust for temporal offsets between slices due to the interleaved acquisition sequence. Spatial normalization was then conducted utilizing Advanced Normalization Tools (ANTs) for precise image registration to a standard anatomical space, enhancing comparability across subjects. Spatial smoothing was applied using FSL’s SUSAN with a 5 mm full-width at half-maximum (FWHM) Gaussian kernel to improve signal-to-noise ratio while preserving spatial specificity.

Brain masks were generated using outputs from FreeSurfer and transformed to the scan space to accurately define the brain boundaries for subsequent analyses. To remove motion-related artifacts, ICA-Based Automatic Removal of Motion Artifacts (ICA-AROMA) was employed, leveraging independent component analysis to identify and exclude noise components associated with head motion. Scaling and temporal filtering were performed for intensity normalization and high-pass filtering, respectively, to remove low-frequency signal drifts.

Scrubbing was implemented to identify and exclude frames with excessive motion, ensuring that only high-quality data contributed to the analyses. Finally, first-level statistical analysis was conducted using FSL’s FILM (FMRIB’s Improved Linear Model) with autocorrelation correction to account for temporal dependencies in the fMRI time series data. This comprehensive preprocessing pipeline enhanced data quality and reliability, facilitating accurate detection of task-related neural activity.

Further details of the in-house preprocessing pipeline, including specific steps and parameters, are provided in the Supplementary Materials.

### 2.5 BOLD Response Analysis

First-level statistical analyses were conducted using the outputs from the preprocessing pipelines. The general linear model (GLM) was applied to the preprocessed data to identify task-related neural activity. Design matrices included task regressors convolved with the canonical double-gamma hemodynamic response function. Confound regressors included motion parameters and scrubbed volumes.

Contrasts of interest were defined to identify the Positive BOLD Response (PBR) and Negative BOLD Response (NBR).

Second-level (group-level) analyses combined data across participants using mixed-effects modeling (FLAME stages 1 and 2). Statistical thresholding was performed using cluster-based correction for multiple comparisons, with a cluster-forming threshold of Z > 2.3 and a corrected cluster significance threshold of p < 0.05.

### 2.6 Functional Connectivity Analysis

To assess whether task-evoked activity reflects brain reconfigurations during different tasks better than functional connectivity, we analyzed functional connectivity during each cognitive task. Functional connectivity was assessed using the Multi-session Hierarchical Bayesian Model (MS-HBM), which allows for the estimation of individual-specific functional networks while accounting for both inter-subject and intra-subject variability (Kong et al., 2019).

The MS-HBM assigns probabilistic labels to each cortical vertex, reflecting the likelihood that a vertex belongs to a particular functional network. For each task, we obtained a parcellation map where each vertex was labeled with a network identity. By focusing on the DMN labels, we created binary maps for each task, where vertices belonging to the DMN were marked as 1, and others as 0. We then calculated the Dice coefficient between these binary DMN maps for each pair of tasks to assess the spatial overlap of the DMN across tasks.

Detailed descriptions of the MS-HBM implementation and functional connectivity analyses are provided in the Supplementary Materials.

### 2.7 Similarity Measures

To assess task specificity and compare activation patterns across tasks, similarity measures were computed using the Dice overlap coefficient. Construct validity was calculated to assess whether activation patterns were more similar within cognitive domains than across domains.

Statistical significance of similarities and construct validity scores was evaluated using permutation tests.

## 3 Results

### 3.1 Task Performance

Participants’ accuracy rates and reaction times varied across tasks (Table 2), reflecting differing cognitive demands. These performance metrics confirmed engagement with the tasks and provided context for interpreting neural activation patterns.

**Table 2.** Performance metrics for both datasets. * Picture Vocabulary in dataset 2 ** Word Memorization in dataset 2 *** In cases where the sum of the mean and standard deviation exceeds 100% for accuracy, this representation does not imply that individual accuracy rates exceed 100%, but rather illustrates the spread of the data around the mean.

| **Task** | **Dataset 1** |  | **Dataset 2** |  |
| --- | --- | --- | --- | --- |
|  | **Accuracy (%)** | **Response time (s)** | **Accuracy (%)** | **Response time (s)** |
| Antonyms | 51.00 ± 19.58 | 6.20 ± 1.37 | 55.65 ± 14.43 | 5.99 ± 1.01 |
| Synonyms | 55.66 ± 21.36 | 6.11 ± 1.36 | 60.82 ± 14.80 | 5.37 ± 1.09 |
| Picture Naming* | - | - | 55.61 ± 16.42 | 5.36 ± 0.82 |
| Matrix Reasoning | 49.14 ± 26.65 | 19.77 ± 9.17 | 68.56 ± 17.52 | 18.37 ± 3.36 |
| Letter Set | 70.54 ± 21.37 | 16.64 ± 5.53 | 82.64 ± 13.52 | 14.72 ± 2.86 |
| Paper Folding | 56.25 ± 25.34 | 17.63 ± 8.26 | 80.23 ± 14.48 | 14.38 ± 3.52 |
| Pattern Comp | 90.47 ± 11.11*** | 1.46 ± 0.23 | 97.00 ± 3.42*** | 1.82 ± 0.31 |
| Letter Comp | 75.95 ± 10.12 | 1.56 ± 0.19 | 86.62 ± 4.27 | 1.91 ± 0.29 |
| Digit Symbol | 88.44 ± 14.67*** | 1.36 ± 0.22 | 94.17 ± 2.57 | 2.37 ± 0.41 |
| Word Order** | 43.94 ± 21.12 | 3.10 ± 0.54 | 81.84 ± 15.91 | 3.31 ± 0.59 |
| Pair Association | 63.61 ± 25.19 | 2.53 ± 0.52 | 83.22 ± 20.53*** | 2.98 ± 0.74 |
| [Logical] Memory | 73.44 ± 15.40 | 4.25 ± 0.83 | 87.40 ± 8.67 | 3.72 ± 0.84 |

#### 3.1.1 Accuracy Rates and Mean Response Times Across Tasks

A one-way ANOVA revealed a significant effect of task on accuracy rates *F*(11, 665) = 61.75, *p* < 0.001, indicating that accuracy differed among the twelve tasks. To further investigate these differences, post hoc comparisons using the Tukey HSD test were conducted (detailed results are provided in Table S3).

The one-way ANOVA also showed a significant effect of task on mean response times *F*(11, 665) = 595.42, *p* < 0.001. Post hoc Tukey HSD tests (Table S4) highlighted significant differences.

Figure S2 illustrates the mean accuracy rates across tasks, and Figure S3 shows the mean response times. Error bars represent the standard error of the mean.

#### 3.1.2 Accuracy Rates Across Cognitive Domains

Grouping tasks by cognitive domain, a one-way ANOVA revealed a significant effect on accuracy rates *F*(3, 673) = 188.96, *p* < 0.001. Post hoc Tukey HSD tests (Table 3) showed that speed of processing tasks had significantly higher accuracy rates than all other domains. The mean difference between speed of processing and fluid reasoning tasks was 15.63% (p < 0.001), and between speed of processing and vocabulary tasks was 35.49% (p < 0.001). Episodic memory tasks had significantly higher accuracy rates than fluid reasoning tasks (mean difference = 6.96%, p < 0.001) and vocabulary tasks (mean difference = 26.82%, p < 0.001). Vocabulary tasks had the lowest accuracy rates, significantly lower than all other domains.

**Table 3.** Post hoc Tukey HSD results for accuracy rates across cognitive domains.

| Comparison | Mean Difference (%) | p-value | 95% CI Lower | 95% CI Upper | Significant |
| --- | --- | --- | --- | --- | --- |
| Episodic Memory vs. Fluid Reasoning | -6.96 | <0.001 | -10.89 | -3.03 | Yes |
| Episodic Memory vs. Speed of Processing | 8.67 | <0.001 | 4.69 | 12.65 | Yes |
| Episodic Memory vs. Vocabulary | -26.82 | <0.001 | -30.84 | -22.8 | Yes |
| Fluid Reasoning vs. Speed of Processing | 15.63 | <0.001 | 11.73 | 19.53 | Yes |
| Fluid Reasoning vs. Vocabulary | -19.86 | <0.001 | -23.8 | -15.91 | Yes |
| Speed of Processing vs. Vocabulary | -35.49 | <0.001 | -39.48 | -31.5 | Yes |

#### 3.1.3 Mean Response Times Across Cognitive Domains

A significant effect of cognitive type on mean response times was also found *F*(3, 673) = 1,665.34, *p* < 0.001. Post hoc Tukey HSD tests (Table 4) indicated that fluid reasoning tasks had significantly longer response times than all other domains. The mean difference compared to episodic memory tasks was 12,493 ms (p < 0.001), and compared to speed of processing tasks was 13,804 ms (p < 0.001). Speed of processing tasks had significantly shorter response times than episodic memory (mean difference = −1,311 ms, p < 0.001) and vocabulary tasks (mean difference = −3,558 ms, p < 0.001).

**Table 4.** Post hoc Tukey HSD results for mean response times across cognitive domains.

| **Comparison** | **Mean Difference (ms)** | **p-value** | **95% CI Lower** | **95% CI Upper** | **Significant** |
| --- | --- | --- | --- | --- | --- |
| Episodic Memory vs. Fluid Reasoning | 12493.35 | <0.001 | 11927.7 | 13059.0 | Yes |
| Episodic Memory vs. Speed of Processing | -1310.72 | <0.001 | -1883.48 | -737.97 | Yes |
| Episodic Memory vs. Vocabulary | 2247.134 | <0.001 | 1668.355 | 2825.913 | Yes |
| Fluid Reasoning vs. Speed of Processing | -13804.1 | <0.001 | -14365.3 | -13242.8 | Yes |
| Fluid Reasoning vs. Vocabulary | -10246.2 | <0.001 | -10813.6 | -9678.79 | Yes |
| Speed of Processing vs. Vocabulary | 3557.858 | <0.001 | 2983.324 | 4132.391 | Yes |

These results indicate that participants performed best and responded fastest on speed of processing tasks, suggesting these tasks were less cognitively demanding or more familiar. Fluid reasoning tasks were the most challenging, reflected in lower accuracy rates and significantly longer response times, indicating higher cognitive load. Vocabulary tasks had the lowest accuracy rates, potentially due to complexity or difficulty in language comprehension, despite moderate response times. Episodic memory tasks showed intermediate performance, with accuracy rates higher than vocabulary and fluid reasoning tasks but lower than speed of processing tasks.

Figure 2.a illustrates the mean accuracy rates across cognitive domains, and Figure 2.b shows the mean response times. Error bars represent the standard error of the mean.

**Figure 2.**
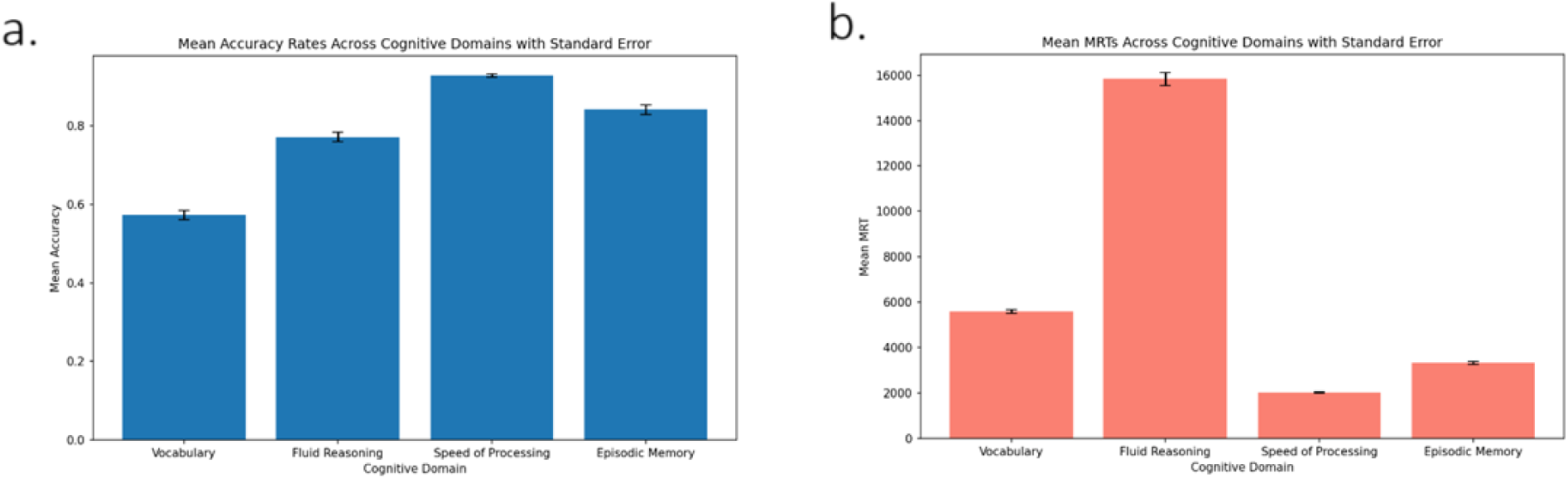
(a) Mean accuracy rates and (b) mean response times across cognitive domains.

### 3.2 Spatial Patterns of BOLD Responses

Group-level activation and deactivation maps were generated for each of the twelve cognitive tasks to examine the spatial distribution of BOLD Responses (PBRs and NBRs). Across all tasks, PBRs were predominantly observed in a consistent set of brain regions associated with cognitive functions. Common areas of activation included the bilateral dorsolateral prefrontal cortex (DLPFC), anterior cingulate cortex (ACC), and superior parietal lobules. These regions are known to be involved in attentional control, working memory, and executive functions. NBRs were primarily observed in regions associated with the DMN, including the medial prefrontal cortex, posterior cingulate cortex, precuneus, and bilateral inferior parietal lobules. The spatial patterns of NBRs exhibited notable variability across different cognitive tasks.

Figure 3 and Figure S4 illustrate the group-level PBR activation maps as well as NBR deactivation maps for datasets 1 and 2, respectively, for representative tasks from each cognitive domain. The figures demonstrate the variability in PBR and NBR patterns across tasks, with deactivations in different DMN regions corresponding to the cognitive demands of each task.

**Figure 3.**
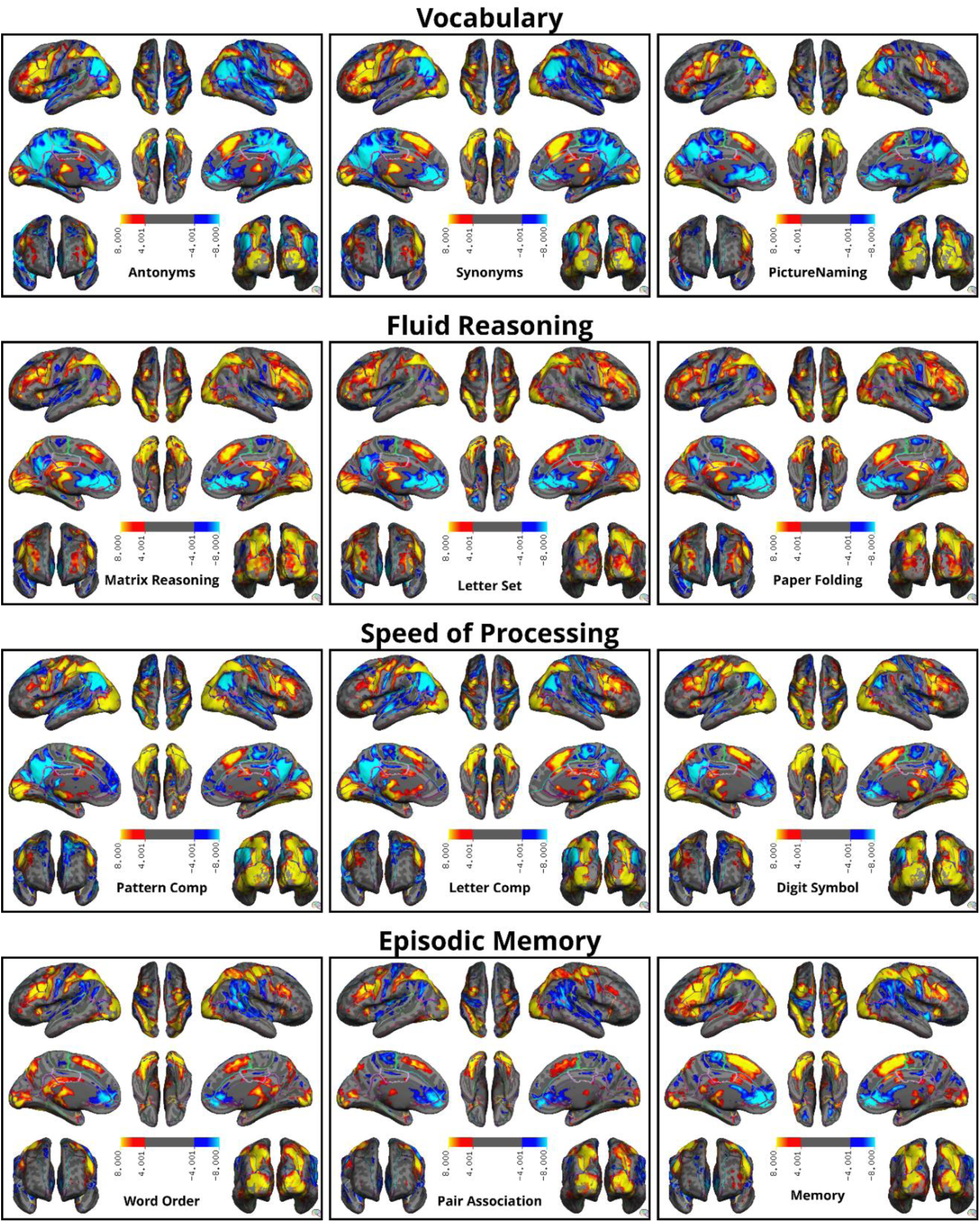
Group-level BOLD Responses (PBR and NBR) activation and deactivation maps for representative tasks for dataset 1. The red and yellow regions represent areas of positive activation, or PBR, while the blue regions indicate deactivation, or NBR. Common regions of activation include the dorsolateral prefrontal cortex, anterior cingulate cortex, and superior parietal lobules. Deactivations are primarily observed in default mode network regions, with variability across tasks. Memory tasks show deactivations in posterior DMN regions, while fluid reasoning tasks exhibit deactivations in anterior DMN regions. Task-specific activations and deactivations are observed in different areas for different cognitive tasks.

#### 3.2.1 Task-Specific Activation and Deactivation Patterns

Detailed analysis revealed that NBR patterns were more variable and task-specific compared to PBRs. For instance, during vocabulary tasks, such as Antonyms and Synonyms, we found that the entire DMN is deactivated to some extent. This widespread deactivation suggests that the brain suppresses internal processes to focus on language comprehension and semantic processing required for vocabulary tasks. The deactivation of self-referential and introspective regions allows for enhanced attention to external linguistic stimuli.

In fluid reasoning tasks like Matrix Reasoning and Paper Folding, an area encompassing the Rostral Anterior Cingulate Cortex is deactivated. This region is involved in emotional regulation. Deactivation here may indicate that fluid reasoning tasks require suppression of emotional processing to enhance logical reasoning capabilities. However, the Inferior Parietal Lobule is not deactivated. This region is associated with attention and spatial processing, which are crucial for solving complex problems and engaging in logical reasoning. Its continued activity suggests that it remains active to support the cognitive demands of fluid reasoning tasks.

For speed of processing tasks, such as Digit Symbol and Letter Comparison, we observed that the Posterior Cingulate and Inferior Parietal Lobule are deactivated, indicating a suppression of self-referential thoughts and shifting of attentional resources to enhance processing speed. Additionally, the Medial Orbitofrontal is not deactivated. This region is involved in reward processing and decision-making. Its sustained activity may facilitate quick decision-making and motivation needed to perform these tasks rapidly.

In episodic memory tasks, like Logical Memory and Pair Association, we also see deactivation in the Rostral Anterior Cingulate Cortex, similar to fluid reasoning tasks. However, the deactivation pattern differs. Additionally, Posterior Cingulate is not deactivated. It plays a significant role in memory retrieval and integration. Its continued activity is essential for successful performance in episodic memory tasks, as it supports the retrieval and processing of memory-related information. The brain maintains activity in this area to facilitate the required memory functions, indicating why it remains active during these tasks.

In dataset 1, comparative analyses using Dice similarity coefficients demonstrated lower spatial overlap among NBR deactivation maps compared to PBR activation maps. The mean Dice overlap coefficient (activation/deactivation maps overlap between each two tasks where 0 indicates no overlap and 1 indicates perfect overlap) for NBR maps between tasks was 0.444 (SD: 0.130), significantly lower than that for PBR (0.668 (SD: 0.085)) with t(65) = 18.375, *p* < .001 (the degree of freedom (df) is n-1 where n is the number of task pairs: 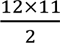).

Similarity matrices constructed for both PBR and NBR showed higher within-domain similarities compared to between-domain similarities for both datasets as is shown in Figure 4. Construct validity score (the average similarity within the same cognitive task types (inside the pink rectangles) minus the average similarity across different task types) for NBR was higher (0.239, *p* < .001) compared to PBR (0.121, *p* < .001), indicating that NBR patterns were more specific to the cognitive domain and task type.

**Figure 4.**
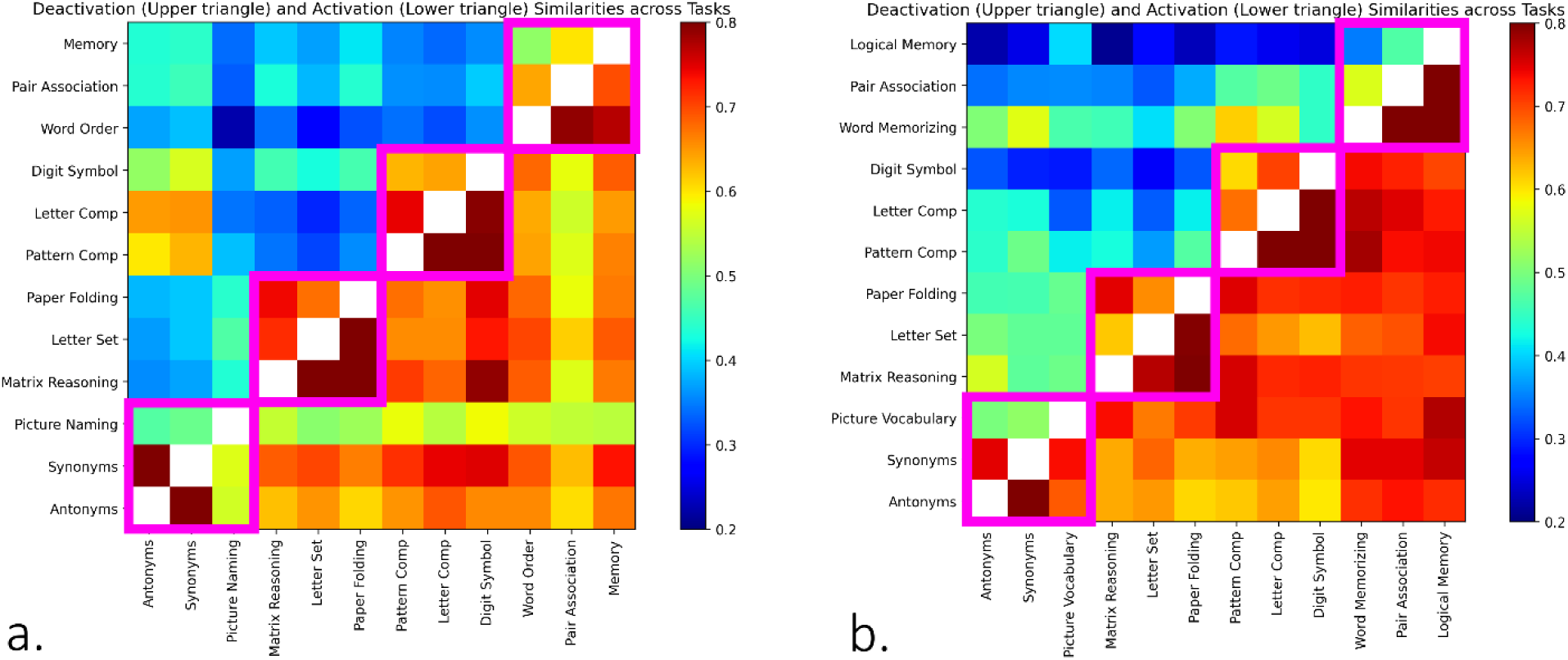
Pair-wise similarities illustrated with color coding matrices created from a) the dataset 1 for activation and deactivation as well as b) the dataset 2 for activation and deactivation patterns for each pair of tasks.

Similar results were observed in dataset 2. The mean Dice coefficient for NBR maps between tasks was 0.437 (SD: 0.127), significantly lower than that for PBR (0.723 (SD: 0.065)) with t(65) = 20.802, *p* < .001. Construct validity score for NBR was higher (0.195, *p* < .001) compared to PBR (0.109, *p* < .001), indicating that NBR patterns were more specific to the cognitive domain and task type.

### 3.3 Functional Connectivity Analysis

#### 3.3.1 Assessment of DMN Parcellation Stability Across Tasks

To determine whether the spatial configuration of major brain networks, specifically the default mode network (DMN), exhibits task-related reconfigurations, we analyzed the consistency of DMN parcellations across the twelve cognitive tasks in Dataset 2. Using the Multi-session Hierarchical Bayesian Model (MS-HBM), we obtained individualized functional parcellations of the cortical surface for each task. The MS-HBM assigns labels (from 1 to 17) to each cortical vertex on the fsaverage surface map, representing different functional networks.

For each task, we focused on the DMN by identifying the vertices labeled as belonging to this network. This resulted in binary maps for each task, where a value of 1 indicated DMN membership, and 0 indicated otherwise. To quantify the spatial overlap of the DMN across tasks, we calculated the Dice similarity coefficient for each pair of tasks.

#### 3.3.2 Stability of DMN Connectivity Across Tasks

The mean Dice coefficient for DMN labels between tasks was 0.777 (SD = 0.027), indicating a high degree of spatial overlap in DMN parcellations across different tasks. The similarity matrix (Figure 5) illustrates the Dice coefficients for all task pairs, showing consistently high values across the matrix.

**Figure 5.**
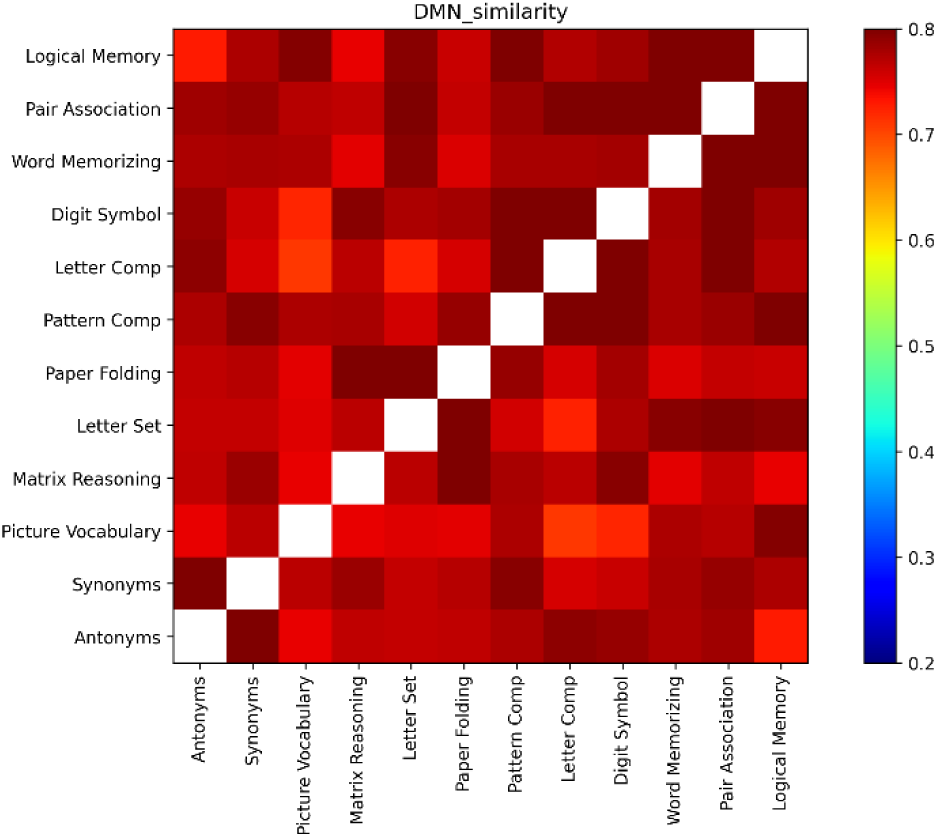
Pair-wise similarities illustrated with a color-coded matrix representing the Dice coefficients of DMN label overlap between each pair of tasks. High Dice coefficients indicate substantial spatial overlap of DMN regions across tasks, suggesting stability in the DMN’s spatial configuration.

To assess whether the DMN parcellations were more similar within cognitive domains than between domains, we calculated the construct validity score by subtracting the average between-domain Dice coefficient from the average within-domain Dice coefficient. The construct validity score was 0.035, *p* < .001, which is close to zero. This suggests that there were no significant differences in DMN spatial configuration between different cognitive domains.

#### 3.3.3 Comparison with Task-Evoked BOLD Responses

These findings indicate that the spatial configuration of the DMN remains largely consistent across different cognitive tasks. In contrast, the NBR and PBR patterns showed greater task specificity, with distinct spatial patterns corresponding to different cognitive demands.

This suggests that while the overall spatial extent of major brain networks like the DMN does not reconfigure substantially with task demands, the localized neural activity within these networks (as reflected by NBR and PBR) exhibits task-specific variations. Therefore, task-evoked BOLD responses may provide more sensitive measures of task-specific neural dynamics than changes in the spatial configuration of functional networks.

## 4 Discussion

This study investigated whether the Negative BOLD Response (NBR) exhibits greater task specificity than the Positive BOLD Response (PBR) across various cognitive tasks and assessed whether task-evoked activity reflects task-induced brain reconfigurations better than functional connectivity. Our findings demonstrate that NBR patterns are significantly more task-specific than PBR patterns, with distinct spatial deactivations corresponding to different cognitive domains. In contrast, functional connectivity of the default mode network (DMN) remains largely stable across tasks, suggesting that task-evoked activity captures task-specific neural dynamics more effectively than functional connectivity measures.

Our results reveal that NBR patterns vary significantly across different cognitive tasks, exhibiting distinct spatial deactivations aligned with the specific cognitive demands. In vocabulary tasks such as Antonyms and Synonyms, widespread deactivation was observed across the DMN, suggesting suppression of self-referential and introspective processes to facilitate language comprehension and semantic processing. This aligns with previous studies indicating that DMN deactivation enhances external attentional focus required for language tasks (Binder et al., 2009)(Humphreys & Lambon Ralph, 2015).

In fluid reasoning tasks like Matrix Reasoning and Paper Folding, significant deactivation occurred in the rostral anterior cingulate cortex (ACC), a region involved in emotional regulation. Deactivation here may indicate the suppression of emotional processing to enhance logical reasoning capabilities (Duncan & Owen, 2000)(Luo et al., 2007). The lack of deactivation in the inferior parietal lobule, associated with attention and spatial processing, suggests its continued engagement during complex problem-solving (Uttal et al., 2013).

Speed of processing tasks, such as Digit Symbol and Letter Comparison, showed deactivations in the posterior cingulate cortex and inferior parietal lobule, indicating suppression of self-referential thoughts and reallocation of attentional resources to enhance processing speed. The sustained activity in the medial orbitofrontal cortex, involved in reward processing and decision-making, may facilitate quick decision-making and motivation required for rapid task performance (Heyes & Foster, 2002)(Rolls, 2000).

In episodic memory tasks like Logical Memory and Pair Association, deactivation was observed in the rostral ACC, similar to fluid reasoning tasks, but not in the posterior cingulate cortex. The posterior cingulate cortex plays a crucial role in memory retrieval and integration (Leech & Sharp, 2014)(Sestieri et al., 2011). Its continued activity underscores its importance in supporting memory-related functions during these tasks.

In contrast, PBR patterns were more generalized across tasks, with common activations in regions such as the dorsolateral prefrontal cortex, ACC, and superior parietal lobules. While some task-specific activations were noted, the overall spatial patterns of PBRs showed substantial overlap between tasks.

These findings suggest that NBRs are more sensitive to the specific cognitive processes engaged by different tasks, reflecting localized neural deactivations that facilitate task performance by suppressing irrelevant neural activity. The enhanced task specificity of NBRs aligns with prior research indicating that NBRs can provide more precise mapping of functional brain areas (Shmuel et al., 2006).

Our functional connectivity analyses revealed that the spatial configuration of the DMN remains stable across different cognitive tasks. The high mean Dice coefficient (0.777 ± 0.027) indicates substantial spatial overlap of DMN regions between tasks. The construct validity score was close to zero, suggesting no significant differences in DMN spatial configuration between cognitive domains.

These results imply that, at the level of network parcellation, the DMN does not undergo significant spatial reconfigurations in response to different cognitive demands. This stability contrasts with the task-specific variations observed in NBR patterns, indicating that task-evoked BOLD responses capture dynamic neural changes not reflected in the overall network configuration.

This finding aligns with previous studies suggesting that while the brain’s intrinsic functional architecture remains relatively consistent, task demands modulate neural activity within networks rather than altering their spatial extent (Cole et al., 2014)(Krienen et al., 2014). Task-evoked activity, particularly NBRs, may thus provide a more sensitive measure of task-specific neural reconfigurations than functional connectivity measures derived from network parcellation.

The greater task specificity of NBRs has significant implications for our understanding of the neural mechanisms underlying cognitive processes. The observed deactivation patterns suggest that the brain actively suppresses certain regions to optimize performance on specific tasks, supporting the notion of neural efficiency. By deactivating non-essential areas, the brain may reduce interference and conserve cognitive resources (Neubauer & Fink, 2009).

For example, widespread DMN deactivation during vocabulary tasks may facilitate focused attention on external linguistic stimuli by reducing internally directed thoughts. Similarly, suppression of the rostral ACC during fluid reasoning and episodic memory tasks may minimize emotional interference, enhancing cognitive control and memory retrieval processes.

These task-specific NBR patterns highlight the importance of considering both activations and deactivations in cognitive neuroscience research. While PBRs indicate regions engaged during task performance, NBRs provide complementary information about regions disengaged to facilitate efficient processing. This dual perspective offers a more comprehensive understanding of the neural dynamics involved in cognitive functions.

The findings also have clinical relevance, particularly in conditions characterized by aberrant DMN activity. Disruptions in DMN deactivation have been associated with cognitive deficits and symptom severity in disorders such as Alzheimer’s disease, depression, and schizophrenia (Broyd et al., 2009)(Whitfield-Gabrieli & Ford, 2012)(Zhang & Raichle, 2010). The enhanced task specificity of NBRs may make them valuable biomarkers for detecting and monitoring such dysfunctions.

Understanding task-specific NBR patterns could inform interventions aimed at modulating neural activity to improve cognitive performance. Techniques such as neurofeedback or non-invasive brain stimulation could potentially target specific regions to enhance or suppress activity, based on patterns identified in healthy individuals.

Despite the strengths of this study, several limitations should be acknowledged. First, the sample consisted of healthy adults under the age of 50, which may limit the generalizability of the findings to older populations or clinical groups. Future studies should include a broader age range and individuals with neurological or psychiatric conditions to examine whether the observed patterns hold in different populations.

Second, our measure of functional connectivity was based on the spatial overlap of network labels derived from MS-HBM parcellation. While this method assesses the stability of network configurations, it does not capture temporal fluctuations or dynamic connectivity changes that may occur during task performance (Calhoun et al., 2014)(Preti et al., 2017). Employing methods that assess dynamic functional connectivity could reveal additional insights into task-specific neural reconfigurations.

Finally, although we observed greater task specificity in NBR patterns, the underlying neural mechanisms of NBRs are not fully understood. NBRs may reflect decreased neural activity, increased inhibition, or vascular factors (Shmuel et al., 2002)(Mullinger et al., 2017). Further research using techniques such as simultaneous EEG-fMRI or neurophysiological recordings could help elucidate the neural correlates of NBRs.

## 5 Conclusion

In conclusion, our study provides evidence that the Negative BOLD Response exhibits greater task specificity than the Positive BOLD Response across a range of cognitive tasks. The distinct spatial patterns of NBRs correspond to different cognitive demands, highlighting their potential as sensitive indicators of task-specific neural activity. In contrast, functional connectivity of the DMN remains stable across tasks, suggesting that task-evoked activity reflects task-induced brain reconfigurations better than functional connectivity measures based on network parcellation.

These findings enhance our understanding of the neural mechanisms underlying cognitive processes and underscore the importance of considering NBRs in cognitive neuroscience research. By acknowledging the role of neural deactivations alongside activations, we can develop more nuanced models of brain function that account for both the engagement and disengagement of neural networks during cognitive tasks.

## Ethical Considerations

All procedures were approved by the Institutional Review Board of Columbia University Irving Medical Center and Weill Cornell Medical Center. Participants provided written informed consent and were compensated for their participation. MRI safety protocols were strictly adhered to, and participants were screened for contraindications. Data confidentiality was maintained through anonymization and secure storage.

## Acknowledgments

We gratefully acknowledge the funding provided, which made this study possible. We also thank the participants for their time and commitment to this research. Additionally, we appreciate the support and resources provided by Columbia University Irving Medical Center and Weill Cornell Medical Center.

## Data availability

The data for this project is confidential but may be obtained with Data Use Agreements with the Weill Cornell Medicine and Columbia University Irving Medical Center. It can take some weeks to negotiate data use agreements and gain access to the data. The author will assist with any reasonable replication attempts for the following publication.

## Supp1. Methods

### 5.1 Task Design and Timing

Tasks were carefully designed to control for extraneous variables and ensure that observed neural activations were attributable to the cognitive processes of interest.

- Stimulus Presentation:

`o` Standardized across participants.
`o` Visual stimuli presented using MRI-compatible display systems.
`o` Auditory stimuli delivered via MRI-compatible headphones.
- Response Recording:

`o` Button boxes used for manual responses.
`o` Verbal responses recorded when necessary, with noise-cancellation measures in place.
- Timing Parameters:

`o` Consistent stimulus durations and inter-trial intervals within each task.
`o` Timing optimized to capture the hemodynamic response associated with each cognitive process.
- Task Order:

`o` Randomized across participants to prevent order effects.
`o` Breaks provided between tasks to reduce fatigue.
- Control Conditions:

`o` Fixation crosses or baseline tasks used during rest periods to establish a baseline for neural activity.

### 5.2 In-House Preprocessing Pipeline for Dataset 2

Detailed steps of the preprocessing pipeline implemented for Dataset 2:

- **TopUp Distortion Correction**: Susceptibility-induced distortions were corrected using FSL’s TopUp tool. An opposite phase-encoding scan was selected based on minimal displacement calculated using the Euclidean norm of differences in coordinates after applying rotation and translation matrices. The selected scan was transformed into the original scan’s space using orientation matrices.
- **Motion Correction and Realignment**: The 4D fMRI data were realigned using FSL’s MCFLIRT, with the single-band reference (SBRef) image as the reference.
- **Slice Timing Correction**: Adjusted for temporal offsets between slices using FSL’s slicetimer tool.
- **Spatial Normalization**: An in-house spatial normalization technique was developed using outputs from TopUp, FreeSurfer, and Advanced Normalization Tools (ANTs). Warping fields were generated to transform images between the scan space, structural space, and MNI152 standard space.
- **Spatial Smoothing**: Applied using FSL’s SUSAN with a full-width at half maximum (FWHM) of 5 mm.
- **Brain Mask Creation**: Created by transforming the brain mask from the subject’s structural space (obtained from FreeSurfer) to the scan space using the inverse warping field.
- **ICA-Based Automatic Removal of Motion Artifacts (ICA-AROMA)**: Employed to identify and remove motion-related artifacts from the fMRI data.
- **Scaling and Temporal Filtering**: The denoised data were intensity-normalized and temporally filtered using a high-pass filter with a cutoff frequency corresponding to 100 seconds.
- **Scrubbing**: Frames with excessive motion were identified using framewise displacement (FD) and excluded from analysis.
- **Parzen Windowing for Task Timing Verification**: A time window was slid over the data by adjusting the stimulation onsets in the task timing file to verify synchronization between the stimuli and the fMRI data. The optimal shift corresponded to maximum activation in the visual cortex.

### 5.3 Functional Connectivity Analysis Details

- **MS-HBM Implementation**: The Multi-session Hierarchical Bayesian Model was used to estimate functional networks at both group and individual levels. The model assumed that vertices within the same network share similar connectivity profiles. Individual-specific parcellations were refined using the Variational Bayes Expectation Maximization algorithm.
- **Network Definition and Identification**: Criteria and thresholds used for identifying the default mode network (DMN), dorsal attention network (DAN), and their sub-networks were established based on prior literature and model estimations.

### 5.4 Similarity Measures and Statistical Analysis

- **Dice Overlap Coefficient**: Calculated as 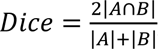, where *A* and *B* are binary activation maps for two tasks.
- **Construct Validity Calculation**: Assessed by comparing the mean similarity of activation patterns within cognitive domains to the mean similarity across domains.
- **Permutation Tests**: Statistical significance was evaluated by randomly shuffling task labels to generate a null distribution of similarity scores.

## Supp2. Results

### 5.5 Task Performance

#### 5.5.1 Accuracy Rates Across Tasks

The post hoc analysis revealed several significant differences in accuracy rates between tasks. Speed of processing tasks, such as Digit Symbol and Pattern Comparison, had significantly higher accuracy rates compared to tasks in other domains. For example, the Digit Symbol task showed a mean accuracy difference of 38.53% compared to the Antonyms task (p < 0.001) and 33.36% compared to the Synonyms task (p < 0.001). Vocabulary tasks, including Antonyms and Synonyms, had lower accuracy rates compared to most other tasks. The Antonyms task had significantly lower accuracy than tasks like Letter Comparison (mean difference = −30.97%, p < 0.001) and Logical Memory (mean difference = −31.76%, p < 0.001). Fluid reasoning tasks, such as Matrix Reasoning and Paper Folding, had moderate accuracy rates but were significantly lower than speed of processing tasks. Matrix Reasoning had a significantly lower accuracy than Pattern Comparison (mean difference = −28.44%, p < 0.001).

These findings indicate that participants performed significantly better on speed of processing tasks compared to vocabulary and fluid reasoning tasks.

#### 5.5.2 Mean Response Times Across Tasks

Fluid reasoning tasks, particularly Matrix Reasoning, had significantly longer response times than all other tasks. Participants took, on average, 16,003 ms longer to complete Matrix Reasoning compared to the Digit Symbol task (p < 0.001) and 12,384 ms longer compared to the Antonyms task (p < 0.001). Speed of processing tasks, such as Digit Symbol and Letter Comparison, had the shortest response times. For instance, the Digit Symbol task was completed 3,619 ms faster than the Antonyms task (p < 0.001). Vocabulary and memory tasks had intermediate response times, significantly shorter than fluid reasoning tasks but longer than speed of processing tasks.

These results suggest that fluid reasoning tasks were more time-consuming, reflecting higher cognitive demand, whereas speed of processing tasks were completed more quickly.

## Supp3. Figures

**Figure S1.**
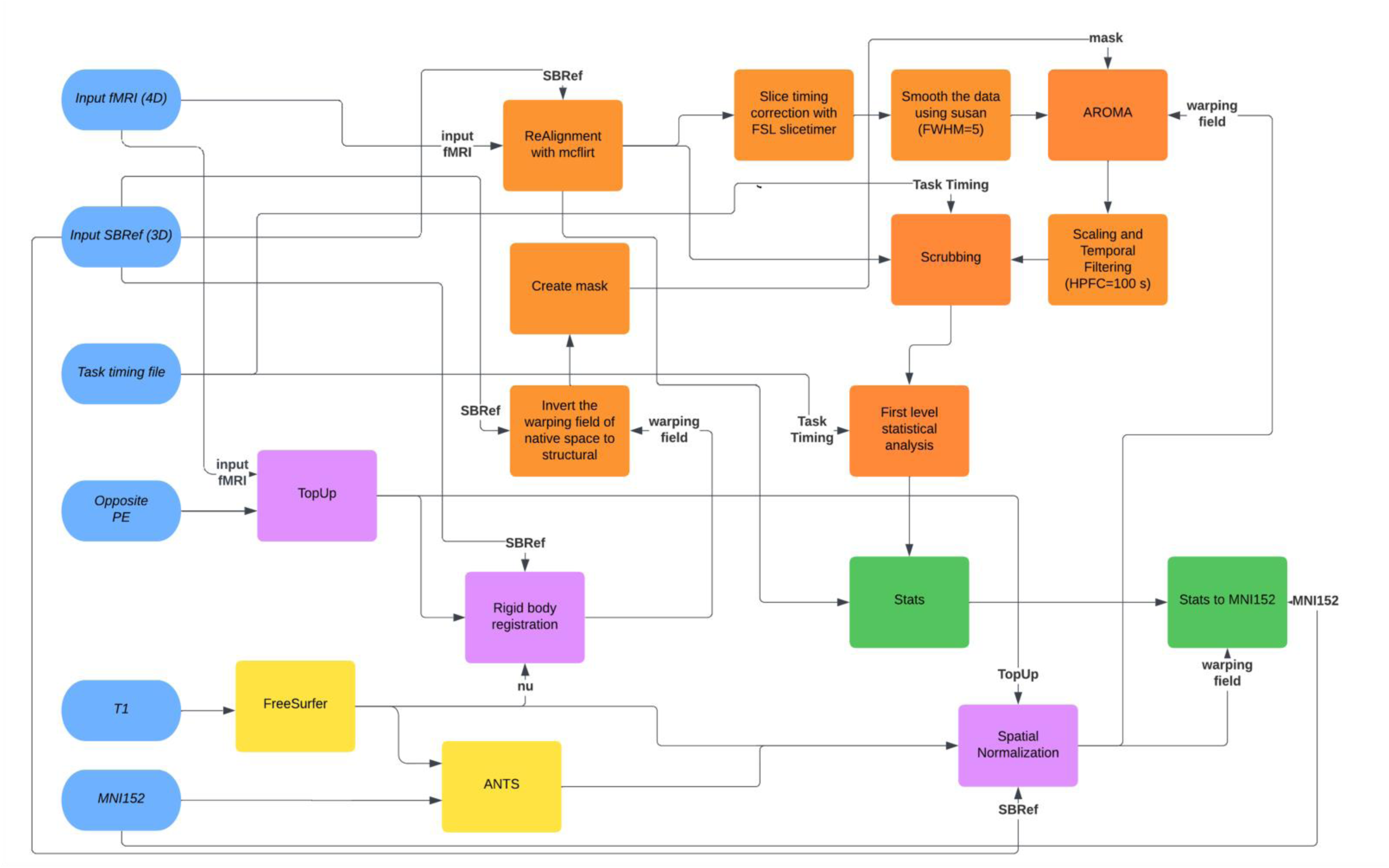
Flowchart of the first-level fMRI data analysis pipeline

**Figure S2.**
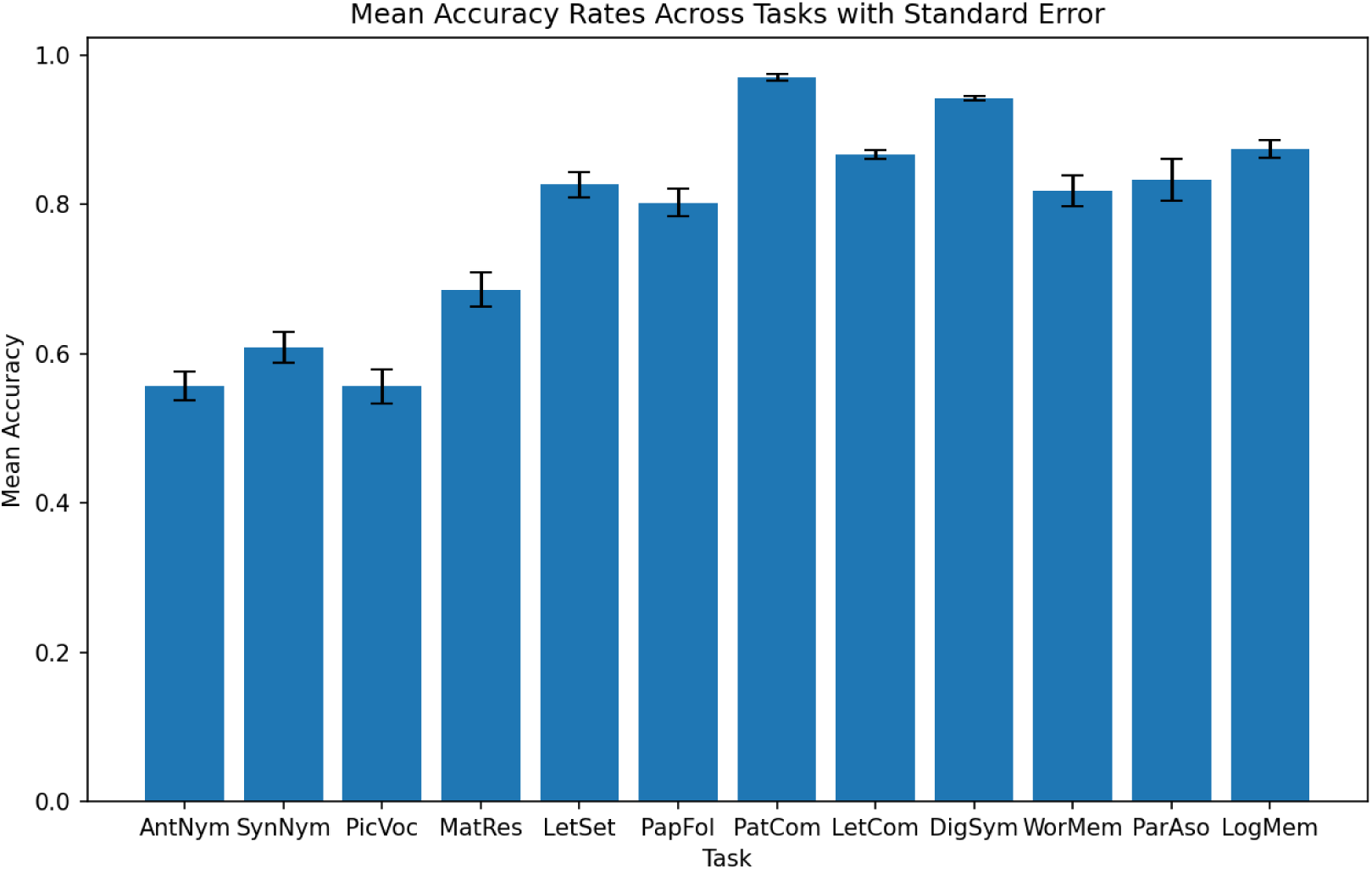
Mean accuracy rates across tasks

**Figure S3.**
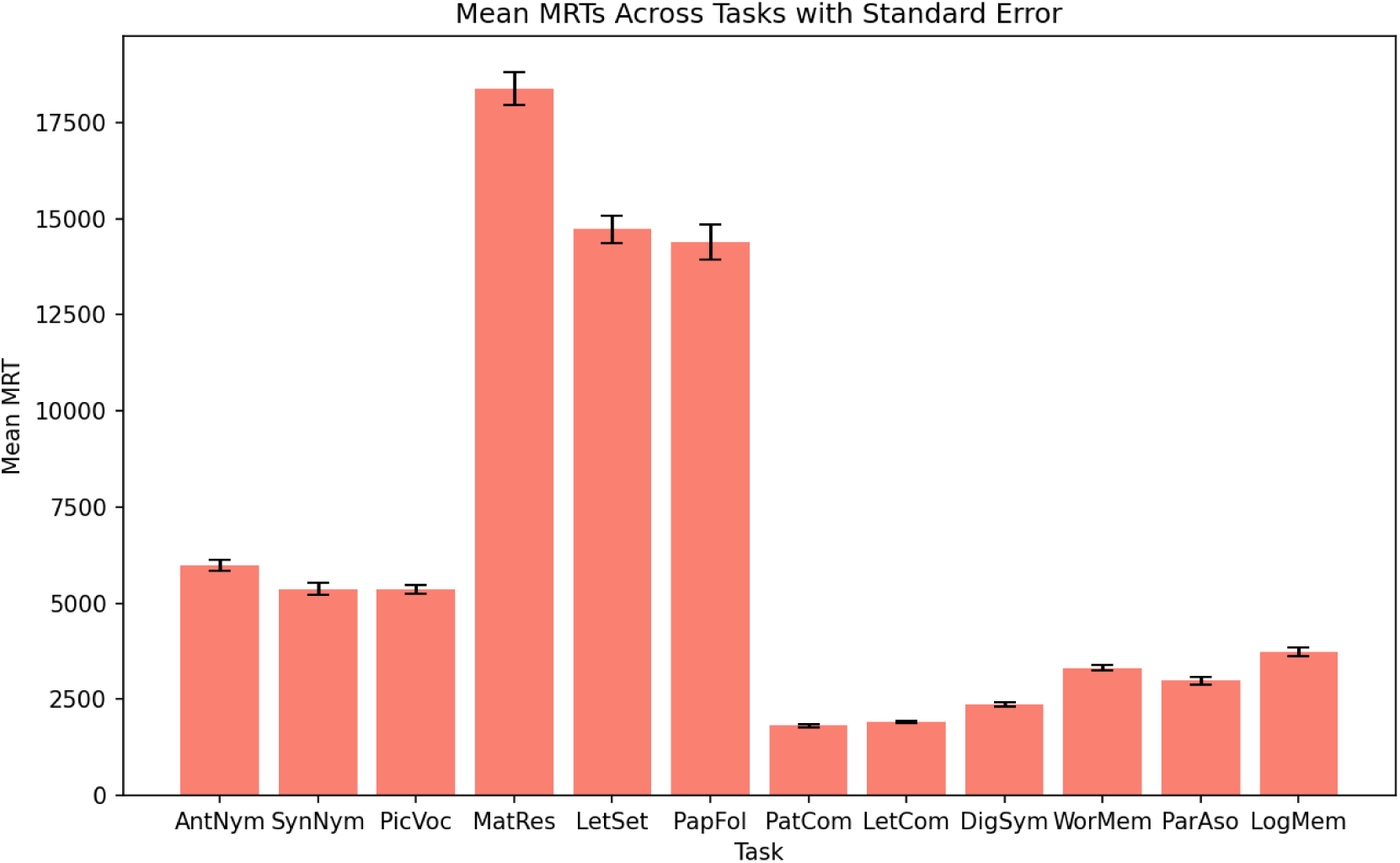
Mean response times across tasks

**Figure S4.**
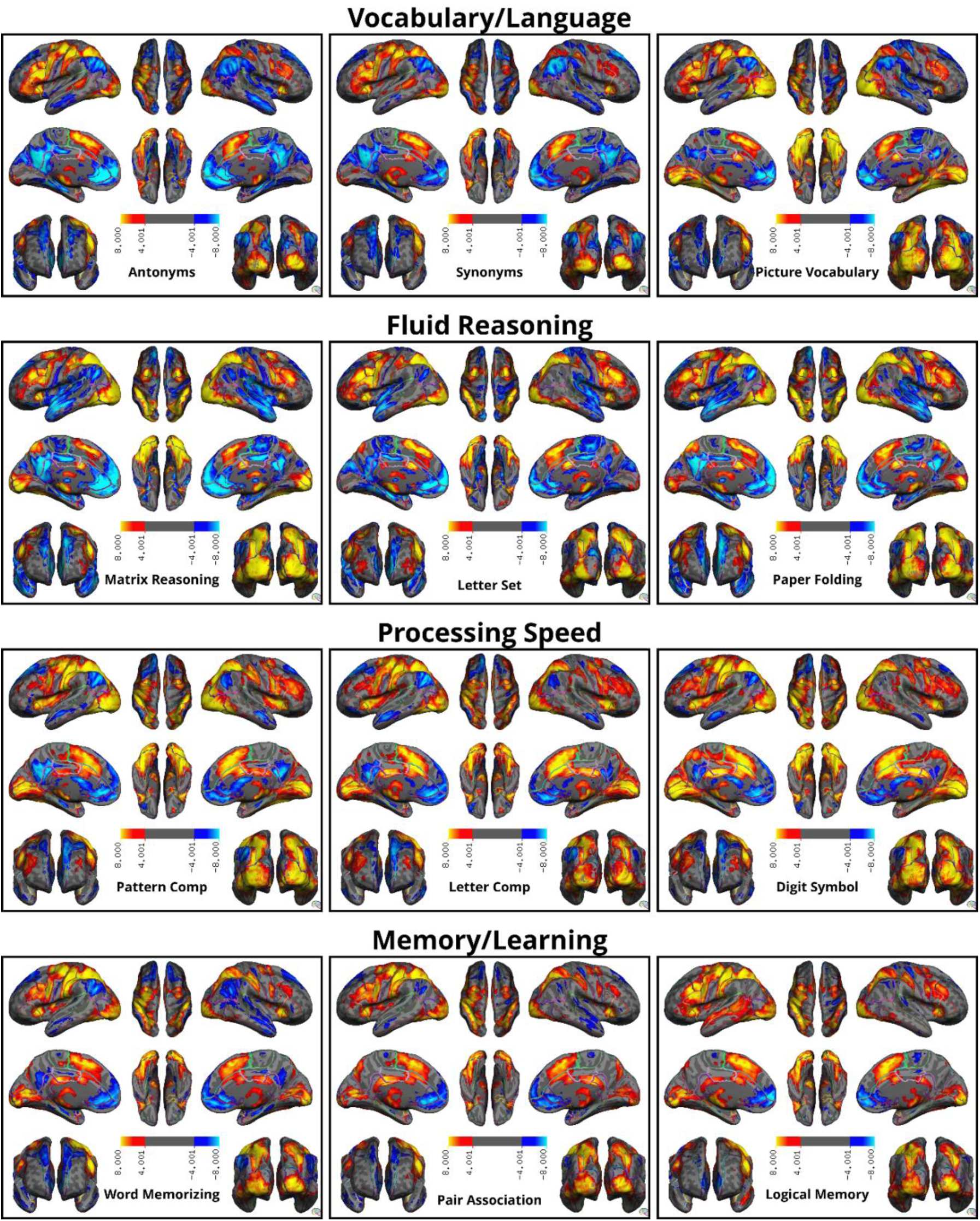
Group-level BOLD Responses (PBR and NBR) activation and deactivation maps for representative tasks for dataset 2. The red and yellow regions represent areas of positive activation, or PBR, while the blue regions indicate deactivation, or NBR. Common regions of activation include the dorsolateral prefrontal cortex, anterior cingulate cortex, and superior parietal lobules. Deactivations are primarily observed in default mode network regions, with variability across tasks. Memory tasks show deactivations in posterior DMN regions, while fluid reasoning tasks exhibit deactivations in anterior DMN regions. Task-specific activations and deactivations are observed in different areas for different cognitive tasks.

## Supp4. Tables

**Table S1.** Task specifics for dataset 1.

| **Task** | **BF(s)** | **EoR** | **NoS** | **NoP** | **TpB** | **TpR** | **LoT(s)** | **LoB** | **LoS** | **LoP** | **ISI(s)** | **ITI** | **IPI** |
| --- | --- | --- | --- | --- | --- | --- | --- | --- | --- | --- | --- | --- | --- |
| Antonyms | 36 | 6' 26" | - | - | 3 | - | 13.5 | 42 | - | - | 0.5 | - | - |
| Synonyms | 36 | 6' 26" | - | - | 3 | - | 13.5 | 42 | - | - | 0.5 | - | - |
| Picture Naming | 36 | 6' 16" | - | - | 8 | - | 4.5 | 40 | - | - | 0.5 | - | - |
| Matrix Reasoning | 24 | 14' 26" | - | - | - | 7~18 | 11~85 | - | - | - | - | 35 | - |
| Letter Set | 24 | 14' 26" | - | - | - | 7~18 | 11~85 | - | - | - | - | 35 | - |
| Paper Folding | 24 | 14' 26" | - | - | - | 7~18 | 11~85 | - | - | - | - | 35 | - |
| Pattern Comp | 36 | 6' 26" | - | - | 12 | - | 3 | 42 | - | - | 0.5 | - | - |
| Letter Comp | 36 | 6' 26" | - | - | 12 | - | 3 | 42 | - | - | 0.5 | - | - |
| Digit Symbol | 36 | 7' 4" | - | - | 18 | - | 2.5 | 49.5 | - | - | 0.25 | - | - |
| Word Order | 30 | 7' 2" | 12 | 10 | - | 2 | - | - | 4 | 6 | 0.7~11.4 | 30 | 2 |
| Pair Association | 30 | 3' 24" | 6 | 6 | - | 2 | - | - | 2 | 5 | 0.2~5.6 | 10 | 5 |
| [Logical] Memory | 30 | 7' 0" | 3 | 10 | - | 2 | - | - | 10 | 10 | 0 | 30 | 2.5 |

**Table S2.**
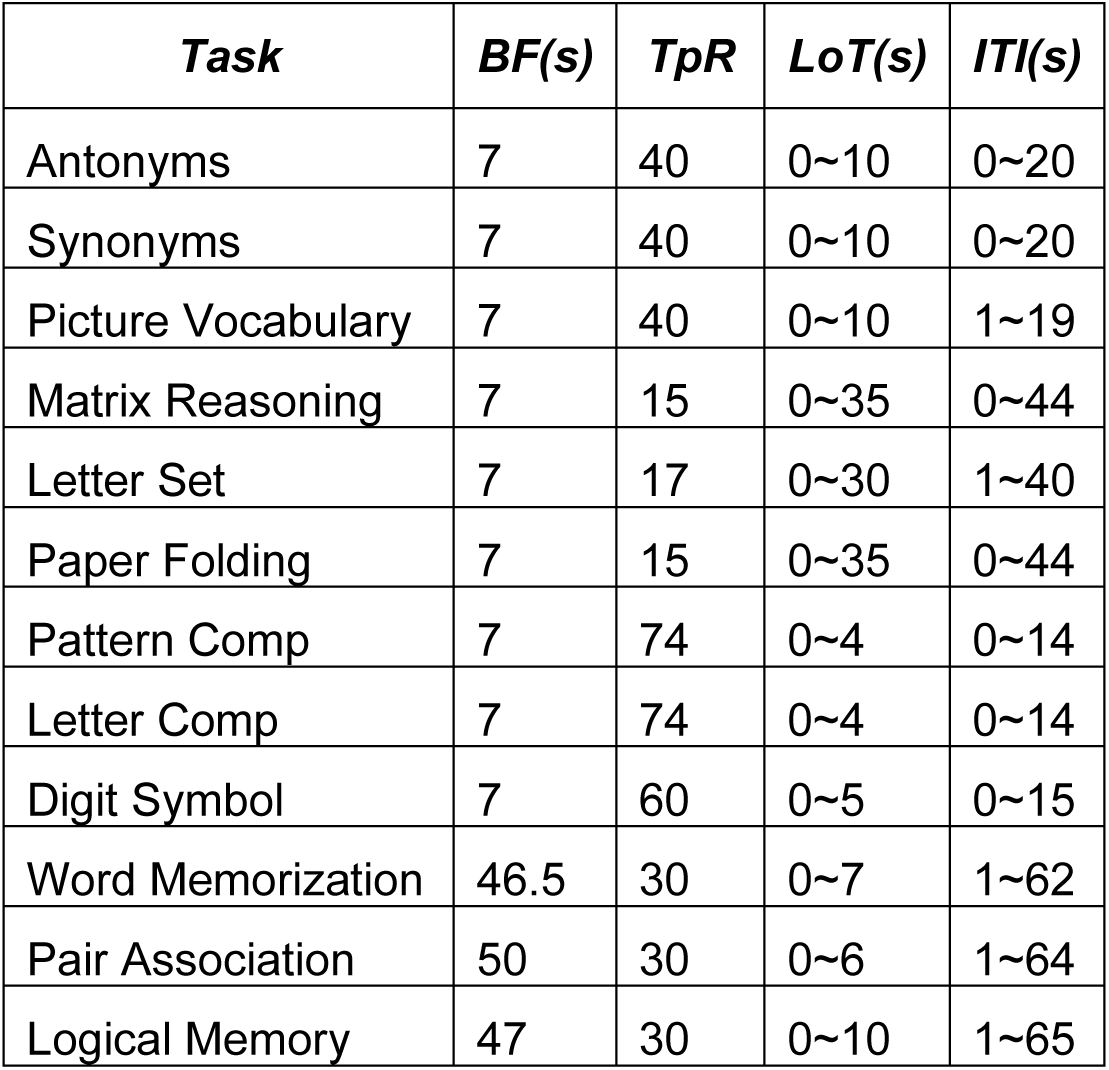
Task specifics for dataset 2.

**Table S3.**
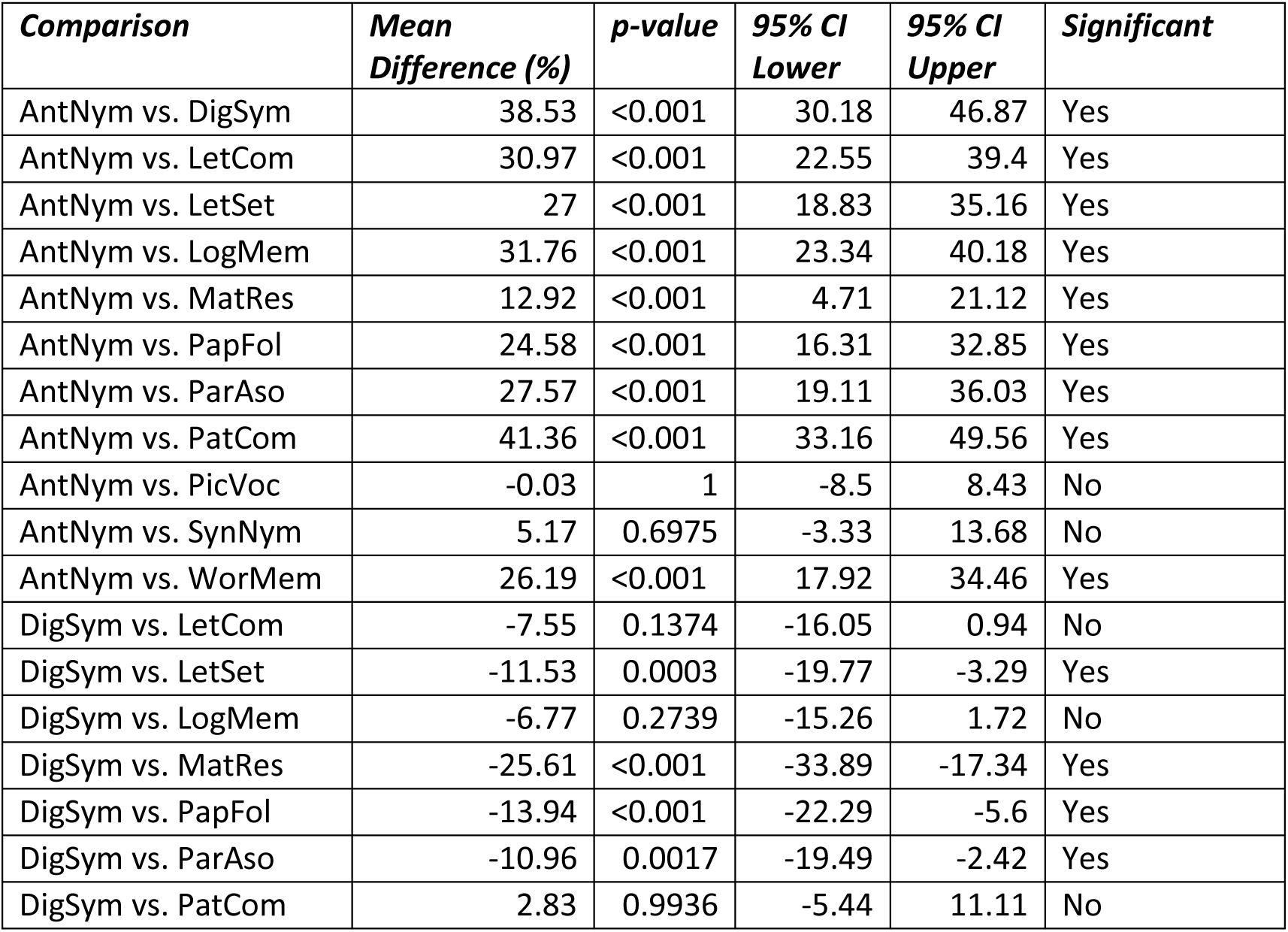
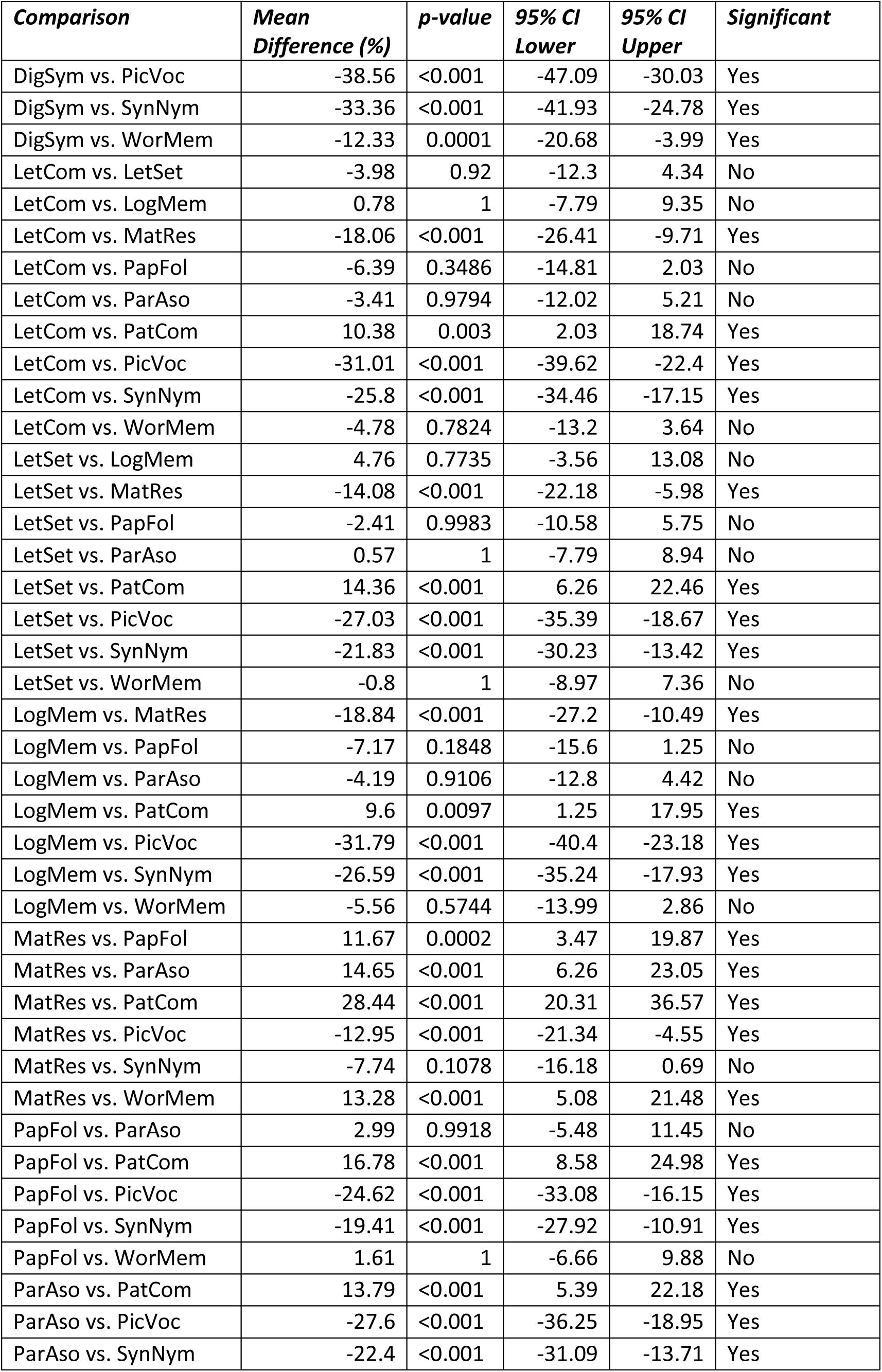
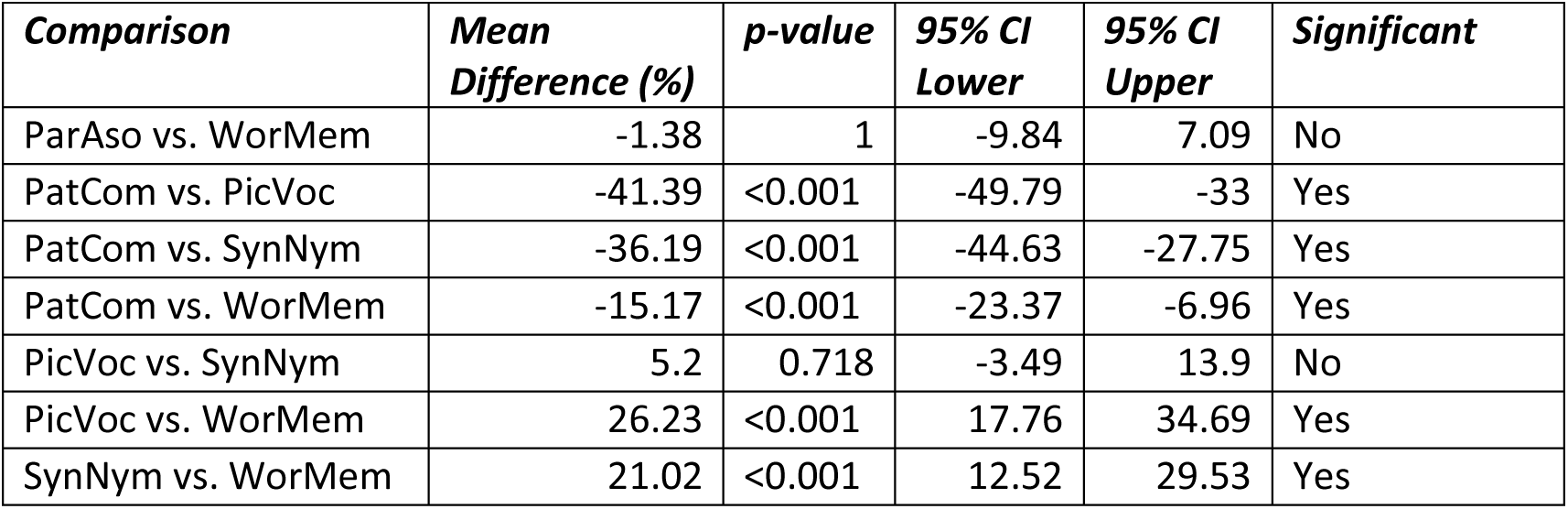
Post hoc Tukey HSD results for accuracy rates across tasks.

| **Comparison** | **Mean Difference (%)** | **p-value** | **95% CI Lower** | **95% CI Upper** | **Significant** |
| --- | --- | --- | --- | --- | --- |
| AntNym vs. DigSym | 38.53 | <0.001 | 30.18 | 46.87 | Yes |
| AntNym vs. LetCom | 30.97 | <0.001 | 22.55 | 39.4 | Yes |
| AntNym vs. LetSet | 27 | <0.001 | 18.83 | 35.16 | Yes |
| AntNym vs. LogMem | 31.76 | <0.001 | 23.34 | 40.18 | Yes |
| AntNym vs. MatRes | 12.92 | <0.001 | 4.71 | 21.12 | Yes |
| AntNym vs. PapFol | 24.58 | <0.001 | 16.31 | 32.85 | Yes |
| AntNym vs. ParAso | 27.57 | <0.001 | 19.11 | 36.03 | Yes |
| AntNym vs. PatCom | 41.36 | <0.001 | 33.16 | 49.56 | Yes |
| AntNym vs. PicVoc | -0.03 | 1 | -8.5 | 8.43 | No |
| AntNym vs. SynNym | 5.17 | 0.6975 | -3.33 | 13.68 | No |
| AntNym vs. WorMem | 26.19 | <0.001 | 17.92 | 34.46 | Yes |
| DigSym vs. LetCom | -7.55 | 0.1374 | -16.05 | 0.94 | No |
| DigSym vs. LetSet | -11.53 | 0.0003 | -19.77 | -3.29 | Yes |
| DigSym vs. LogMem | -6.77 | 0.2739 | -15.26 | 1.72 | No |
| DigSym vs. MatRes | -25.61 | <0.001 | -33.89 | -17.34 | Yes |
| DigSym vs. PapFol | -13.94 | <0.001 | -22.29 | -5.6 | Yes |
| DigSym vs. ParAso | -10.96 | 0.0017 | -19.49 | -2.42 | Yes |
| DigSym vs. PatCom | 2.83 | 0.9936 | -5.44 | 11.11 | No |

| **Comparison** | **Mean**<br>**Difference (%)** | **p-value** | **95% CI**<br>**Lower** | **95% CI**<br>**Upper** | **Significant** |
| --- | --- | --- | --- | --- | --- |
| DigSym vs. PicVoc | -38.56 | <0.001 | -47.09 | -30.03 | Yes |
| DigSym vs. SynNym | -33.36 | <0.001 | -41.93 | -24.78 | Yes |
| DigSym vs. WorMem | -12.33 | 0.0001 | -20.68 | -3.99 | Yes |
| LetCom vs. LetSet | -3.98 | 0.92 | -12.3 | 4.34 | No |
| LetCom vs. LogMem | 0.78 | 1 | -7.79 | 9.35 | No |
| LetCom vs. MatRes | -18.06 | <0.001 | -26.41 | -9.71 | Yes |
| LetCom vs. PapFol | -6.39 | 0.3486 | -14.81 | 2.03 | No |
| LetCom vs. ParAso | -3.41 | 0.9794 | -12.02 | 5.21 | No |
| LetCom vs. PatCom | 10.38 | 0.003 | 2.03 | 18.74 | Yes |
| LetCom vs. PicVoc | -31.01 | <0.001 | -39.62 | -22.4 | Yes |
| LetCom vs. SynNym | -25.8 | <0.001 | -34.46 | -17.15 | Yes |
| LetCom vs. WorMem | -4.78 | 0.7824 | -13.2 | 3.64 | No |
| LetSet vs. LogMem | 4.76 | 0.7735 | -3.56 | 13.08 | No |
| LetSet vs. MatRes | -14.08 | <0.001 | -22.18 | -5.98 | Yes |
| LetSet vs. PapFol | -2.41 | 0.9983 | -10.58 | 5.75 | No |
| LetSet vs. ParAso | 0.57 | 1 | -7.79 | 8.94 | No |
| LetSet vs. PatCom | 14.36 | <0.001 | 6.26 | 22.46 | Yes |
| LetSet vs. PicVoc | -27.03 | <0.001 | -35.39 | -18.67 | Yes |
| LetSet vs. SynNym | -21.83 | <0.001 | -30.23 | -13.42 | Yes |
| LetSet vs. WorMem | -0.8 | 1 | -8.97 | 7.36 | No |
| LogMem vs. MatRes | -18.84 | <0.001 | -27.2 | -10.49 | Yes |
| LogMem vs. PapFol | -7.17 | 0.1848 | -15.6 | 1.25 | No |
| LogMem vs. ParAso | -4.19 | 0.9106 | -12.8 | 4.42 | No |
| LogMem vs. PatCom | 9.6 | 0.0097 | 1.25 | 17.95 | Yes |
| LogMem vs. PicVoc | -31.79 | <0.001 | -40.4 | -23.18 | Yes |
| LogMem vs. SynNym | -26.59 | <0.001 | -35.24 | -17.93 | Yes |
| LogMem vs. WorMem | -5.56 | 0.5744 | -13.99 | 2.86 | No |
| MatRes vs. PapFol | 11.67 | 0.0002 | 3.47 | 19.87 | Yes |
| MatRes vs. ParAso | 14.65 | <0.001 | 6.26 | 23.05 | Yes |
| MatRes vs. PatCom | 28.44 | <0.001 | 20.31 | 36.57 | Yes |
| MatRes vs. PicVoc | -12.95 | <0.001 | -21.34 | -4.55 | Yes |
| MatRes vs. SynNym | -7.74 | 0.1078 | -16.18 | 0.69 | No |
| MatRes vs. WorMem | 13.28 | <0.001 | 5.08 | 21.48 | Yes |
| PapFol vs. ParAso | 2.99 | 0.9918 | -5.48 | 11.45 | No |
| PapFol vs. PatCom | 16.78 | <0.001 | 8.58 | 24.98 | Yes |
| PapFol vs. PicVoc | -24.62 | <0.001 | -33.08 | -16.15 | Yes |
| PapFol vs. SynNym | -19.41 | <0.001 | -27.92 | -10.91 | Yes |
| PapFol vs. WorMem | 1.61 | 1 | -6.66 | 9.88 | No |
| ParAso vs. PatCom | 13.79 | <0.001 | 5.39 | 22.18 | Yes |
| ParAso vs. PicVoc | -27.6 | <0.001 | -36.25 | -18.95 | Yes |
| ParAso vs. SynNym | -22.4 | <0.001 | -31.09 | -13.71 | Yes |
| ParAso vs. WorMem | -1.38 | 1 | -9.84 | 7.09 | No |
| PatCom vs. PicVoc | -41.39 | <0.001 | -49.79 | -33 | Yes |
| PatCom vs. SynNym | -36.19 | <0.001 | -44.63 | -27.75 | Yes |
| PatCom vs. WorMem | -15.17 | <0.001 | -23.37 | -6.96 | Yes |
| PicVoc vs. SynNym | 5.2 | 0.718 | -3.49 | 13.9 | No |
| PicVoc vs. WorMem | 26.23 | <0.001 | 17.76 | 34.69 | Yes |
| SynNym vs. WorMem | 21.02 | <0.001 | 12.52 | 29.53 | Yes |

**Table S4.** Post hoc Tukey HSD results for mean response times across tasks.

| **Comparison** | **Mean Difference (ms)** | **p-value** | **95% CI Lower** | **95% CI Upper** | **Significant** |
| --- | --- | --- | --- | --- | --- |
| AntNym vs. DigSym | -3618.7112 | <0.001 | -4727.098 | -2510.3244 | Yes |
| AntNym vs. LetCom | -4076.6232 | <0.001 | -5195.4042 | -2957.8423 | Yes |
| AntNym vs. LetSet | 8729.7335 | <0.001 | 7644.7041 | 9814.7629 | Yes |
| AntNym vs. LogMem | -2267.8825 | <0.001 | -3386.6635 | -1149.1015 | Yes |
| AntNym vs. MatRes | 12384.0471 | <0.001 | 11294.6197 | 13473.4746 | Yes |
| AntNym vs. PapFol | 8396.5 | <0.001 | 7297.8789 | 9495.1211 | Yes |
| AntNym vs. ParAso | -3003.3692 | <0.001 | -4127.6027 | -1879.1358 | Yes |
| AntNym vs. PatCom | -4166.2195 | <0.001 | -5255.647 | -3076.7921 | Yes |
| AntNym vs. PicVoc | -621.7466 | 0.8101 | -1745.98 | 502.4869 | No |
| AntNym vs. SynNym | -618.7208 | 0.82 | -1748.5886 | 511.1469 | No |
| AntNym vs. WorMem | -2673.4138 | <0.001 | -3772.0349 | -1574.7927 | Yes |
| DigSym vs. LetCom | -457.912 | 0.9749 | -1586.2843 | 670.4602 | No |
| DigSym vs. LetSet | 12348.4447 | <0.001 | 11253.5283 | 13443.361 | Yes |
| DigSym vs. LogMem | 1350.8287 | 0.0054 | 222.4565 | 2479.2009 | Yes |
| DigSym vs. MatRes | 16002.7583 | <0.001 | 14903.4835 | 17102.0332 | Yes |
| DigSym vs. PapFol | 12015.2112 | <0.001 | 10906.8244 | 13123.598 | Yes |
| DigSym vs. ParAso | 615.342 | 0.8285 | -518.4366 | 1749.1205 | No |
| DigSym vs. PatCom | -547.5083 | 0.8962 | -1646.7832 | 551.7665 | No |
| DigSym vs. PicVoc | 2996.9646 | <0.001 | 1863.1861 | 4130.7432 | Yes |
| DigSym vs. SynNym | 2999.9904 | <0.001 | 1860.6247 | 4139.3561 | Yes |
| DigSym vs. WorMem | 945.2974 | 0.1835 | -163.0894 | 2053.6842 | No |
| LetCom vs. LetSet | 12806.3567 | <0.001 | 11700.9195 | 13911.7939 | Yes |
| LetCom vs. LogMem | 1808.7407 | <0.001 | 670.1568 | 2947.3247 | Yes |
| LetCom vs. MatRes | 16460.6704 | <0.001 | 15350.916 | 17570.4247 | Yes |
| LetCom vs. PapFol | 12473.1232 | <0.001 | 11354.3423 | 13591.9042 | Yes |
| LetCom vs. ParAso | 1073.254 | 0.09 | -70.688 | 2217.196 | No |
| LetCom vs. PatCom | -89.5963 | 1 | -1199.3507 | 1020.1581 | No |
| LetCom vs. PicVoc | 3454.8767 | <0.001 | 2310.9346 | 4598.8187 | Yes |

| **Comparison** | **Mean**<br>**Difference**<br>**(ms)** | **p-**<br>**value** | **95% CI**<br>**Lower** | **95% CI**<br>**Upper** | **Significant** |
| --- | --- | --- | --- | --- | --- |
| LetCom vs. SynNym | 3457.9024 | <0.001 | 2308.4227 | 4607.3822 | Yes |
| LetCom vs. WorMem | 1403.2095 | 0.0026 | 284.4285 | 2521.9904 | Yes |
| LetSet vs. LogMem | -10997.616 | <0.001 | -12103.053 | -9892.1788 | Yes |
| LetSet vs. MatRes | 3654.3137 | <0.001 | 2578.5941 | 4730.0333 | Yes |
| LetSet vs. PapFol | -333.2335 | 0.9975 | -1418.2629 | 751.7959 | No |
| LetSet vs. ParAso | -11733.1027 | <0.001 | -12844.058 | -10622.148 | Yes |
| LetSet vs. PatCom | -12895.953 | <0.001 | -13971.673 | -11820.233 | Yes |
| LetSet vs. PicVoc | -9351.48 | <0.001 | -10462.435 | -8240.5249 | Yes |
| LetSet vs. SynNym | -9348.4543 | <0.001 | -10465.110 | -8231.7979 | Yes |
| LetSet vs. WorMem | -11403.1473 | <0.001 | -12488.177 | -10318.118 | Yes |
| LogMem vs. MatRes | 14651.9296 | <0.001 | 13542.1753 | 15761.684 | Yes |
| LogMem vs. PapFol | 10664.3825 | <0.001 | 9545.6015 | 11783.1635 | Yes |
| LogMem vs. ParAso | -735.4867 | 0.6167 | -1879.4287 | 408.4553 | No |
| LogMem vs. PatCom | -1898.337 | <0.001 | -3008.0914 | -788.5827 | Yes |
| LogMem vs. PicVoc | 1646.1359 | 0.0002 | 502.1939 | 2790.0779 | Yes |
| LogMem vs. SynNym | 1649.1617 | 0.0002 | 499.6819 | 2798.6414 | Yes |
| LogMem vs. WorMem | -405.5313 | 0.9897 | -1524.3123 | 713.2497 | No |
| MatRes vs. PapFol | -3987.5471 | <0.001 | -5076.9746 | -2898.1197 | Yes |
| MatRes vs. ParAso | -15387.4164 | <0.001 | -16502.667 | -14272.165 | Yes |
| MatRes vs. PatCom | -16550.2667 | <0.001 | -17630.422 | -15470.111 | Yes |
| MatRes vs. PicVoc | -13005.7937 | <0.001 | -14121.045 | -11890.543 | Yes |
| MatRes vs. SynNym | -13002.7679 | <0.001 | -14123.698 | -11881.838 | Yes |
| MatRes vs. WorMem | -15057.4609 | <0.001 | -16146.888 | -13968.034 | Yes |
| PapFol vs. ParAso | -11399.8692 | <0.001 | -12524.103 | -10275.636 | Yes |
| PapFol vs. PatCom | -12562.7195 | <0.001 | -13652.147 | -11473.292 | Yes |
| PapFol vs. PicVoc | -9018.2466 | <0.001 | -10142.48 | -7894.0131 | Yes |
| PapFol vs. SynNym | -9015.2208 | <0.001 | -10145.089 | -7885.3531 | Yes |
| PapFol vs. WorMem | -11069.9138 | <0.001 | -12168.535 | -9971.2927 | Yes |
| ParAso vs. PatCom | -1162.8503 | 0.0322 | -2278.1013 | -47.5994 | Yes |
| ParAso vs. PicVoc | 2381.6226 | <0.001 | 1232.3475 | 3530.8978 | Yes |
| ParAso vs. SynNym | 2384.6484 | <0.001 | 1229.8611 | 3539.4357 | Yes |
| ParAso vs. WorMem | 329.9554 | 0.9984 | -794.278 | 1454.1889 | No |
| PatCom vs. PicVoc | 3544.473 | <0.001 | 2429.222 | 4659.7239 | Yes |
| PatCom vs. SynNym | 3547.4987 | <0.001 | 2426.5683 | 4668.4291 | Yes |
| PatCom vs. WorMem | 1492.8057 | 0.0005 | 403.3783 | 2582.2332 | Yes |
| PicVoc vs. SynNym | 3.0258 | 1 | -1151.7615 | 1157.813 | No |
| PicVoc vs. WorMem | -2051.6672 | <0.001 | -3175.9006 | -927.4338 | Yes |
| SynNym vs. WorMem | -2054.693 | <0.001 | -3184.5607 | -924.8252 | Yes |

